# Serological thresholds of risk reduction for infant group B streptococcus disease

**DOI:** 10.64898/2026.05.29.26353453

**Authors:** Liberty Cantrell, Kostas Karampatsas, Nick Andrews, Simon Beach, Emily Bentley, Alberto Berardi, Merijn W Bijlsma, Cemal Cagil Kocana, Olwenn Daniel, Neil French, Tom Hall, Alane Izu, Asma Khalil, Gaurav Kwatra, Mary Kyohere, Shabir A. Madhi, Robert Mboizi, Francesca Miselli, Maryke Nielsen, Natasha Thorn, Diederik van de Beek, Kate F. Walker, Paul T. Heath, Kirsty Le Doare, Merryn Voysey, the PREPARE WP3 Study Group

## Abstract

Vaccines to prevent infant group B streptococcus (GBS) disease are advancing, with licensure likely based on safety and immunologic endpoints rather than clinical efficacy data. This approach requires robust, generalisable serological thresholds of risk reduction (SToRRs). We combined data from six case-control studies in Europe and Africa to define SToRRs for early-onset (EOD) and late-onset (LOD) GBS disease. Across diverse epidemiological and healthcare settings, anti-capsular polysaccharide IgG concentrations were consistently higher in infants who remained disease free than in those who developed disease. Higher antibody concentrations were required to reduce the risk of EOD than LOD, and higher concentrations were required for serotype Ia than for serotype III. This study provides a quantitative framework to support correlates-based evaluation and potential licensure of maternal GBS vaccines.

## Introduction

Group B streptococcus (GBS) causes invasive bacterial disease in over 390,000 infants worldwide annually, including almost 92,000 infant deaths^1^ and causes more infant deaths than HIV^2^. Global incidence of infant invasive GBS (iGBS) disease is approximately 0.49 cases per 1000 live births, ranging from 0.21 per 1000 live births in Southeast Asia to 2.00 per 1000 live births in Southern Africa^3^. Infant iGBS disease can be early-onset disease (EOD), developing on days 0-6 of life, or late-onset disease (LOD), developing on days 7-89 of life.

The only strategy to prevent infant iGBS disease relies on administering intrapartum antibiotic prophylaxis (IAP) to pregnant women who are identified as colonised or at increased risk. IAP screening policies vary with some countries, such as the US, using universal microbiological testing of pregnant women, some countries, such as the UK, using a risk factor-based approach to screening, and others with limited or no IAP policies^4^. IAP prevents EOD but has little to no impact on LOD^5,6^. The World Health Organisation (WHO) recommends universal GBS testing or a risk-based approach for IAP use^7^, however logistical challenges mean these are seldom implemented in low-resource settings. There is concern that IAP policies may contribute to antimicrobial resistance^8–11^. IAP must be administered with sufficient time before birth to provide protection, limiting potential effectiveness during a rapid labour^12,13^. These limitations of IAP mean an alternative approach is needed to prevent infant iGBS disease.

WHO considers GBS vaccine development a global priority to prevent infant iGBS disease^14^. Although GBS is a leading cause of neonatal infections, incidence remains low, meaning a phase III vaccine efficacy trial may require enrolment of at least 40-60,000 pregnant women if infant iGBS disease was the primary endpoint in a high incidence setting (≥2 per 1000 livebirths)^15^. Given the costs and challenges associated with conducting such a large study, licensure using immunological endpoints associated with iGBS disease risk reduction is a proposed pathway, with vaccine effectiveness confirmed post-licensure^14^. A similar approach was successfully used to license vaccines against meningococcal B and C in the UK^16–18^ and meningococcal A in Africa^19^. Vaccine licensure using immunologic data without clinical efficacy data requires evidence that the vaccine induces an immune response strong enough to protect against invasive disease across diverse populations and geographical regions. The amount of antibody corresponding to a particular reduction (e.g. 80%) in risk of disease is a serological threshold of risk reduction (SToRR).

A number of vaccines are in development, and have completed phase 2 trials in pregnant women, including a protein-subunit vaccine (GBS-NN/NN2) and a capsular polysaccharide-protein conjugate vaccine (GBS6)^20^ for which a phase 3 trial has started^21^. These vaccines are intended for administration to pregnant women to protect infants via transplacentally transferred antibody, a strategy that has been successful for a number of other pathogens including tetanus, pertussis, influenza, COVID-19 and RSV^22–27^.

GBS has ten serotypes (Ia, Ib, II-IX), of which six (Ia, Ib, II-V) cause 98% of disease^3^ and are included in the GBS6 vaccine. Naturally-derived serotype-specific immunoglobulin G (IgG) concentrations in maternal, cord and infant blood samples vary by serotype^28,29^. Recent SToRR estimates for six serotypes in the US were broken down by disease onset (EOD/LOD)^30^. Previously, SToRRs for serotypes Ia and III were estimated in Finland and South Africa^28,29,31^.

## Results

We analysed data from six case-control studies in the UK, the Netherlands, Italy, Uganda, Malawi and South Africa (Table S1) to estimate SToRRs for infant iGBS disease. Full study methods are described in the supplementary materials. The combination of studies enabled comparison of natural immunity across diverse settings with one of the largest sample sizes to date, allowing SToRRs to be estimated by serotype and timing of disease onset and compared to recent estimates from the US^30^. To our knowledge, this is the first study to estimate SToRRs for infant iGBS disease across both high- and low-income countries with different IAP strategies.

### Demographics and serotype distribution

Across six countries, 488 iGBS cases (infants aged <90 days who developed iGBS disease) had antibody data available at birth (cord blood, n=28, 5.7%), soon after iGBS disease onset (acute blood, n=442, 91%), or both (n=18, 3.7%). 1004 controls (infants born to colonised mothers but without iGBS disease in the first 90 days of life) had cord blood antibody data available (Table S2). Infants were aged 0-87 days (median 1 day) at iGBS disease onset, with 277 (58%) EOD cases and 201 (42%) LOD cases. The proportion of EOD ranged from 46% to 77%. Age at disease onset was unknown for 10 infants (Table S2).

15-42% of cases were born preterm (<37 weeks gestational age) compared with 1.5-30% of controls. IAP exposure was high among controls, particularly in the UK and Italy where 58% and 86% of control mothers received IAP respectively, with low rates elsewhere (0-16%) (Table S2), though eligibility criteria for controls differed in some countries (Table S1).

Among cases, serotype III was most common followed by serotype Ia. Serotype III caused 271/488 (56%) cases of infant disease, of which 124/271 (46%) was EOD and 143/271 (53%) was LOD. Serotype Ia caused 84/488 (17%) cases, of which 58/84 (69%) was EOD and 25/84 (30%) was LOD. The remaining cases were caused by serotypes Ib, II, IV and V (7.8%, 6.4%, 4.7% and 8.4% respectively) (Table S2). The control group consisted of 1004 infants whose mother was colonised with GBS during pregnancy. 379/1004 (38%) controls were serotype III and 213/1004 (21%) were serotype Ia. The remaining controls were serotypes Ib, II, IV and V (5.6%, 13%, 5.9%, 16% respectively).

A small number of cases (46/488) had cord blood antibody data available for analysis. The remaining 442 (91%) had an acute blood sample collected at the time of disease. Acute blood samples were collected as soon as possible after disease diagnosis (median 4 days after onset, range 0-37 days). Most acute samples (82%) were collected within 7 days of disease onset and included in the primary analyses (Table S2).

### Higher anti-CPS IgG concentrations reduced the risk of iGBS disease

For serotypes Ia and III serotype-specific anti-capsular polysaccharide (CPS) IgG geometric mean concentrations (GMC) were higher in controls than EOD cases (serotype Ia 0.169μg/ml (95% confidence interval (CI) 0.105, 0.271) vs 0.022μg/ml (0.01, 0.049); serotype III 0.089μg/ml (0.069, 0.115) vs 0.014μg/ml (0.01, 0.019) respectively). Other serotypes exhibited the same trend but with overlapping CIs. GMCs for LOD were lower than EOD for serotypes III, IV and V (Figure 1, Table 1). When summarised by country, the general pattern remained the same despite some variability in anti-CPS IgG concentrations across the six countries (Figure S2). There was no clear difference in anti-CPS IgG concentrations for control infants whose mothers received IAP compared with those who did not (Figure S3).

**Fig. 1.**
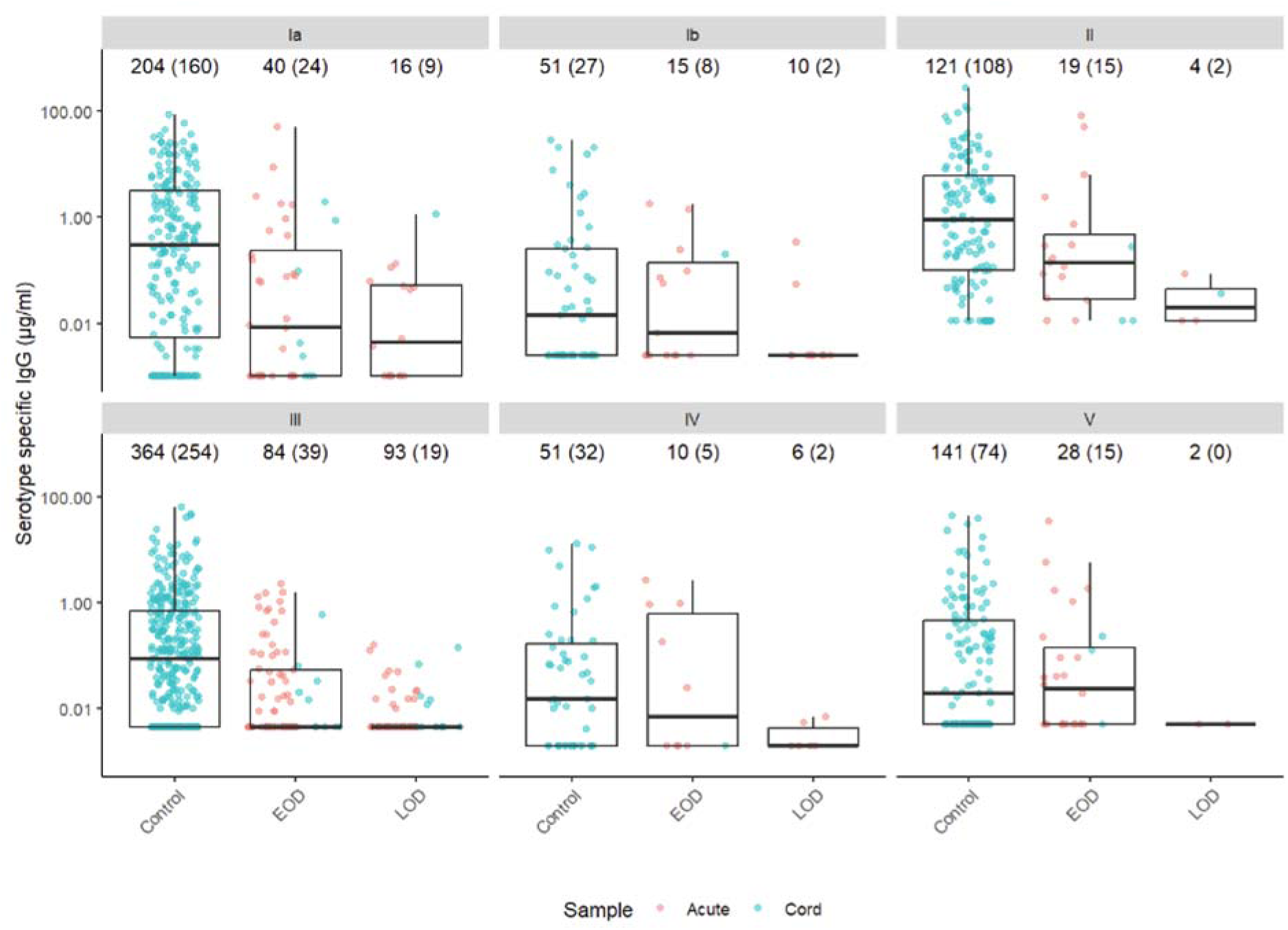
Serotype-specific anti-capsular (CPS) IgG concentrations (_μ_g/ml) in controls, early-onset disease (EOD; 0-6 days) cases and late-onset disease (LOD; 7-89 days) cases, on a log10 scale. Points show individual anti-capsular IgG concentrations, blue points show the antibody concentrations in cord blood samples, red points show antibody measured in acute blood samples. Boxes show the median, upper and lower quartiles. The number of data points is annotated above each boxplot with the number above the LLOQ in brackets.

**Table 1.**
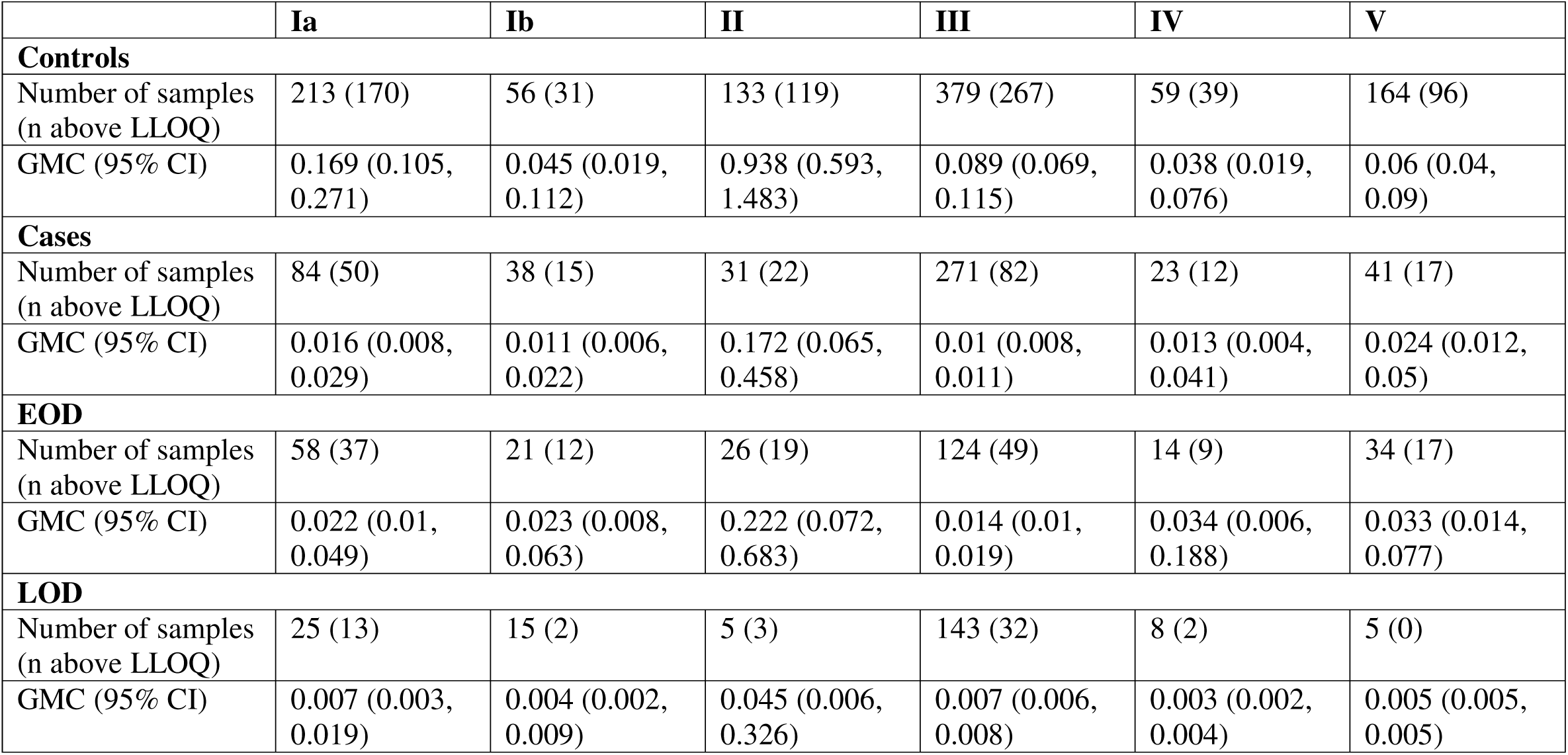
Serotype-specific anti-capsular IgG concentrations (μg/ml) in cases and controls. Geometric mean concentrations (GMC) and 95% confidence intervals (CI) are presented by serotype for controls, cases, cases of early-onset disease (EOD; 0-6 days) and cases of late-onset disease (LOD; 7-89 days). The number of samples and number (n) above the assay lower limit of quantification (LLOQ) are also shown.

|  | **Ia** | **Ib** | **II** | **III** | **IV** | **V** |
| --- | --- | --- | --- | --- | --- | --- |
| **Controls** |  |  |  |  |  |  |
| Number of samples (n above LLOQ) | 213 (170) | 56 (31) | 133 (119) | 379 (267) | 59 (39) | 164 (96) |
| GMC (95% CI) | 0.169 (0.105, 0.271) | 0.045 (0.019, 0.112) | 0.938 (0.593, 1.483) | 0.089 (0.069, 0.115) | 0.038 (0.019, 0.076) | 0.06 (0.04, 0.09) |
| **Cases** |  |  |  |  |  |  |
| Number of samples (n above LLOQ) | 84 (50) | 38 (15) | 31 (22) | 271 (82) | 23 (12) | 41 (17) |
| GMC (95% CI) | 0.016 (0.008, 0.029) | 0.011 (0.006, 0.022) | 0.172 (0.065, 0.458) | 0.01 (0.008, 0.011) | 0.013 (0.004, 0.041) | 0.024 (0.012, 0.05) |
| **EOD** |  |  |  |  |  |  |
| Number of samples (n above LLOQ) | 58 (37) | 21 (12) | 26 (19) | 124 (49) | 14 (9) | 34 (17) |
| GMC (95% CI) | 0.022 (0.01, 0.049) | 0.023 (0.008, 0.063) | 0.222 (0.072, 0.683) | 0.014 (0.01, 0.019) | 0.034 (0.006, 0.188) | 0.033 (0.014, 0.077) |
| **LOD** |  |  |  |  |  |  |
| Number of samples (n above LLOQ) | 25 (13) | 15 (2) | 5 (3) | 143 (32) | 8 (2) | 5 (0) |
| GMC (95% CI) | 0.007 (0.003, 0.019) | 0.004 (0.002, 0.009) | 0.045 (0.006, 0.326) | 0.007 (0.006, 0.008) | 0.003 (0.002, 0.004) | 0.005 (0.005, 0.005) |

The covariate adjusted logit model (CALM)^32^ was used to generate relative risk curves and SToRRs by serotype and disease onset. Full details are given in the Supplementary materials. Higher antibody levels in EOD cases than LOD cases meant higher antibody concentrations were associated with reduced risk of EOD than LOD. For serotype III EOD, the SToRR (95% CI) was 0.662μg/ml (0.234, 1.879) for an 80% risk reduction compared with 0.074μg/ml (0.040, 0.137) for LOD (Table 2). Relative risk curves for serotype Ia were flatter than those for serotype III (Figure 2), resulting in higher anti-CPS concentrations associated with an 80% reduction in relative risk for serotype Ia EOD (5.186μg/ml) and LOD (0.766μg/ml) (Table 2).

**Fig. 2.**
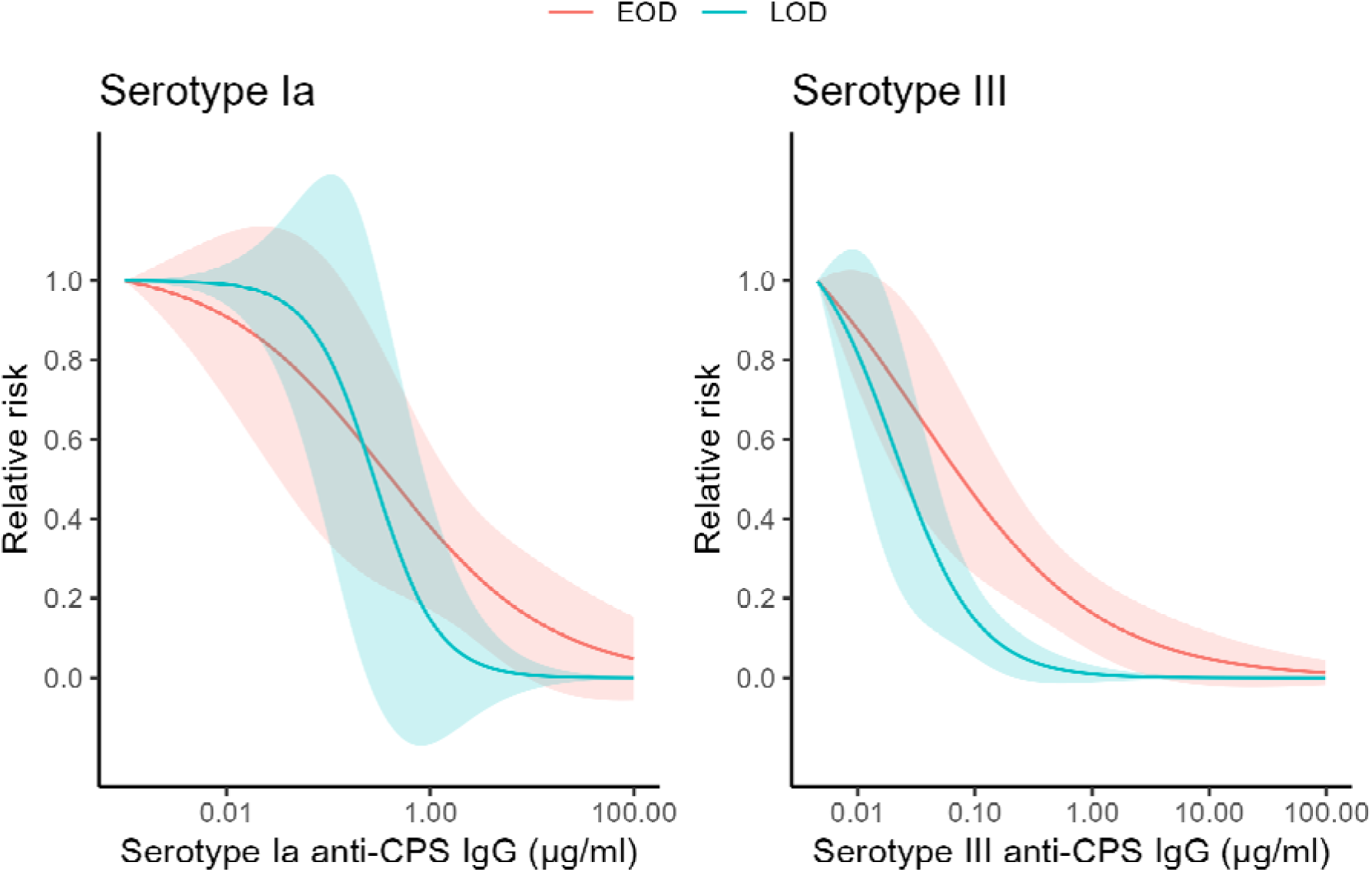
Relative risk curves from the primary CALM analysis for serotypes Ia and III. The solid lines show the relative risk curves, and the shaded regions are the 95% confidence intervals. Relative risk curves for early-onset disease (EOD; 0-6 days) are shown in red, late-onset disease (LOD; 7-89 days) in blue, and the combination of EOD and LOD in black.

**Table 2.** Serotype-specific anti-capsular IgG concentrations (μg/ml) serologic thresholds of risk reduction (SToRR). Table presents the SToRRs and 95% confidence intervals (CI) for 75%, 80% and 90% reductions in relative risk for early-onset disease (EOD; 0-6 days), late-onset disease (LOD; 7-89 days), and for all disease (EOD and LOD) for serotypes Ia and III. SToRRs and 95% CIs were estimated using the covariate adjusted logit model (CALM) and included an adjustment for country. * LOD model not adjusted for country due to low case numbers.

| Serotype | Onset | 75% SToRR (95% CI) | 80% SToRR (95% CI) | 90% SToRR (95% CI) |
| --- | --- | --- | --- | --- |
| III | EOD | 0.415 (0.157, 1.096) | 0.662 (0.234, 1.879) | 2.562 (0.493, 13.323) |
|  | LOD | 0.059 (0.031, 0.113) | 0.074 (0.040, 0.137) | 0.142 (0.071, 0.283) |
|  | All | 0.092 (0.047, 0.180) | 0.138 (0.073, 0.260) | 0.458 (0.220, 0.955) |
| Ia | EOD | 3.056 (0.610, 5.298) | 5.186 (0.823, 32.669) | 23.193 (1.227, 438.548) |
|  | LOD* | 0.623 (0.127, 3.055) | 0.766 (0.145, 4.062) | 1.372 (0.184, 10.220) |
|  | All | 1.646 (0.549, 4.929) | 2.512 (0.783, 8.058) | 8.299 (1.602, 42.996) |

### Sensitivity analyses

#### Analysis method

To test the impact of the choice of statistical method on overall conclusions, analyses were repeated using a Weibull distribution fitted within a Bayesian framework, as used elsewhere^28,29,31^.

For serotype III, the Weibull SToRRs were lower than the CALM SToRRs for EOD and LOD. The 80% Weibull SToRR for serotype III EOD was 0.388μg/ml compared with 0.662μg/ml from the primary CALM analysis, and for LOD the 80% SToRRs from the Weibull and CALM analyses were 0.036μg/ml and 0.074μg/ml respectively. However, for serotype Ia EOD the 80% Weibull SToRR was higher than the CALM SToRR (7.062μg/ml compared with 5.186μg/ml respectively). The 80% Weibull SToRR for serotype Ia LOD was 0.242μg/ml, lower than the 80% CALM SToRR of 0.766μg/ml (Table S3).

#### Acute sample timing

The primary analysis included samples collected within 7 days of disease onset. As a sensitivity analysis, additional samples collected more than 7 days after disease onset were included. For serotypes Ia and III, the 95% CIs for the relative risk curves and SToRRs including additional samples overlapped the 95% CI for the primary analysis. Restricting the sampling window to within 3 days after disease onset removed up to 52% of cases from the analysis, reducing the sample size and increasing width of CIs (Tables S4-5 and Figure S5).

#### LOD controls

Approximately 50% of LOD cases are born to women colonised with the same serotype^33^. To reflect this scenario in the primary analysis, 50% of controls were selected from those exposed to the same serotype as the LOD cases with the remaining controls selected from mothers colonised with a different serotype during pregnancy. As a sensitivity analysis, all controls exposed to the same serotype as LOD cases were selected (100% serotype matched). For serotypes Ia and III, the 95% CIs for relative risk curves using 100% serotype matched controls overlapped the 95% CIs for the primary analysis using 50% serotype matched controls (Table S4 and Figure S6).

#### Antibody waning

Controls and a small number of cases provided cord blood samples. The remaining cases provided acute blood samples at the time of disease. Antibody concentrations measured in acute blood samples are lower than those measured in cord blood samples due to antibody waning after birth, particularly for LOD where there has been more time since birth for antibody waning. We performed a sensitivity analysis to investigate the impact of antibody waning in acute samples on the analysis. Accounting for antibody waning for serotypes Ia and III LOD did not significantly impact the resultant relative risk curve, where the 95% CI for the adjusted analysis overlapped the 95% CI for the primary analysis (Table S4 and Figure S7).

### Predicting vaccine efficacy

Once antibody-risk curves were defined for the major serotypes we estimated predicted vaccine efficacy (pVE) for GBS6 vaccine. We used published phase 2 immunogenicity data^28^ to simulate antibody concentrations in vaccine and placebo recipients for 1000 simulated trials. Antibody risk curves from the current study were used to assign probability of disease and disease status to simulated trial participants. Full details are in the supplementary materials.

Across 1000 simulated vaccine trials, median pVE (95% CI) for serotypes III and Ia EOD were 75% (59%, 86%) and 83% (69%, 93%) respectively. Predicted VE was higher for LOD compared with EOD, with median pVE (95% CI) of 91% (78%, 98%) for serotype III, and 100% (86%, 100%) for serotype Ia (Table S5).

## Discussion

In this large multicountry analysis, we defined SToRRs for infant iGBS disease in high- and low-income settings. Across six geographically and epidemiologically diverse settings, anti-CPS IgG concentrations were higher in infants who remained iGBS disease free than in those who developed iGBS disease, as seen previously^29–31^. Antibody concentrations were higher in cases of EOD than LOD, in line with previous studies^29–31^. This pattern of higher antibody concentrations in EOD cases is reflected in the SToRRs, with higher antibody concentrations associated with reduced risk of EOD. The consistency of these patterns across countries with differing healthcare systems, IAP policies, and baseline disease incidence supports the broader applicability of these findings.

We selected CALM for the primary analysis because it allows adjustment for key sources of heterogeneity inherent to multi-country observational data. This approach yields more conservative SToRR estimates than alternative methods^34^. While conservative thresholds may be preferable from a regulatory perspective, overly stringent SToRRs could inadvertently impede the development of effective vaccines. To address this, we complemented the primary analysis with Bayesian Weibull sensitivity analyses which demonstrated similar overall patterns, providing reassurance that the main conclusions are robust to analytical approach.

We used the Group B Streptococcus: Standardization of Laboratory Assays (GASTON) standardised and validated assay to measure anti-CPS IgG concentrations, enabling meaningful comparison of thresholds across studies. Our estimates are broadly consistent with SToRRs from Finland and the US^30,31^. In Finland, cord blood samples were used exclusively, whereas the US study used dried blood spots, and our analysis included cord blood and acute samples. The Bayesian Weibull method was used to estimate SToRRs in Finland^31^, whereas the US study used the CALM method and a sensitivity analysis using the Bayesian Weibull method which showed the same trends as the CALM analysis^30^. Case-control matching and analytical frameworks also differed. Nonetheless, all studies demonstrated higher antibody concentrations associated with protection against serotype Ia compared with serotype III, and higher concentrations required for protection against EOD than LOD. These convergent findings across independent datasets strengthen confidence in the relevance of the identified thresholds.

It is biologically plausible that higher antibody concentrations are required to reduce the risk of EOD compared with LOD due to potentially different forces of infection. Infants who develop iGBS disease during the first days of life are exposed to GBS over multiple days in utero or during delivery, likely resulting in a higher dose of pathogen exposure than infants exposed to GBS postnatally. This differing exposure intensity may require higher antibody concentrations to prevent invasion during the immediate perinatal period. These differences in exposure levels and the antibody concentration required for protection highlight the importance of distinguishing between EOD and LOD when defining SToRRs.

Serotypes Ia and III were most common among cases, enabling reliable estimates of SToRRs. Differences between serotypes were evident, with consistently higher thresholds for serotype Ia than serotype III. These differences align with prior observations of serotype-specific variability in immunogenicity and disease risk^30^. Together, these results highlight the need for serotype- and onset-specific considerations in the interpretation of immunogenicity data from maternal GBS vaccine trials, rather than reliance on a single universal threshold.

Antibody patterns were consistent across countries, despite substantial heterogeneity in IAP use, prematurity rates, and healthcare contexts. This supports the generalisability of shared SToRRs across diverse settings, rather than requiring country-specific thresholds. Nonetheless, continued validation in additional populations remains important, particularly as vaccine programs expand into new regions and epidemiological contexts.

Using the derived risk curves and published phase 2 immunogenicity data, we predicted vaccine efficacy for a hexavalent GBS conjugate vaccine. We predicted vaccine efficacy for EOD and LOD separately for serotypes Ia and III. Predicted efficacy estimates were high, with lower bounds of the 95% CIs above 50%. Predicted efficacy was higher for LOD than EOD, consistent with lower antibody concentrations required for protection later in infancy. Disease-risk curves from Finland^31^ also predicted high vaccine efficacy for serotype III and all six serotypes combined, using a composite threshold. These estimates of predicted efficacy are indicative of potential efficacy rather than direct substitutes for clinical effectiveness data. Importantly, the modelling assumes that vaccine-induced antibodies confer protection comparable to naturally acquired antibodies, an assumption that underpins correlates-based licensure pathways for several existing vaccines and is widely accepted in regulatory frameworks.

A highly efficacious vaccine against both EOD and LOD could protect a large number of infants from invasive disease, including reducing the number of infants with bacterial meningitis and contributing to WHO’s goal to defeat meningitis by 2030^35^. LOD represents approximately half of all GBS cases, yet current prevention strategies have no impact on LOD. Maternal immunisation offers the first targeted approach to prevent LOD.

Our study has limitations. Our data had low numbers of serotype Ib and II cases, which prevented risk curves from converging for EOD and LOD separately for these rarer serotypes. This issue is not unique in our data. The US study also had fewer cases of the rarer serotypes resulting in step-functions rather than relative risk curves. We saw higher rates of IAP use among controls mothers than case mothers. IAP effectively prevents EOD, so some controls may have developed EOD in the absence of IAP. However, iGBS incidence is less than 1 per 1000 livebirths for EOD and LOD combined, making it unlikely that any of the controls whose mothers received IAP would have developed EOD if IAP had not been administered. Practical challenges meant cord blood samples were not available for most cases in our data, with acute blood samples collected at the time of disease instead. To limit the potential impact of antibody waning between birth and acute sample collection we only included samples collected within 7 days of disease onset in the primary analyses. Sensitivity analyses using different sampling timeframes showed consistent patterns.

A higher proportion of cases were preterm than controls. This relative imbalance prevented case-control matching by gestational age. The protective efficacy of the vaccine in preterm infants remains uncertain, despite this group being particularly vulnerable to LOD. In preterm infants, maternal-foetal antibody transfer is inversely related to gestational age^23,36^. Given that SToRRs are lower for LOD than for EOD, it is plausible that some preterm infants derive some benefit from maternal vaccination. This may help inform optimal timing of maternal immunization during pregnancy.

This study provides robust, multi-country evidence defining serological thresholds associated with reduced risk of infant iGBS disease, and providing important insight into the potential efficacy of vaccines currently in development. Consistent antibody-risk relationships across diverse settings support the use of shared SToRRs to inform the development, evaluation, and licensure of maternal GBS vaccines. By providing a quantitative framework linking antibody concentrations to disease risk reduction, these findings address a critical barrier to vaccine implementation and contribute to global efforts to reduce the burden of neonatal bacterial infections. Availability of large harmonised datasets from high- and low-income settings, aligned through standardised assays and statistical methods, provide confidence that vaccines inducing antibody levels meeting these thresholds have real potential to save lives.

## Methods

### Study description

We combined data from case-control studies conducted in the UK (192 cases, 271 controls), the Netherlands (115 cases, 62 controls), Italy (67 cases, 330 controls), Uganda (24 cases, 71 controls), Malawi (13 cases, 20 controls) and South Africa (77 cases, 250 controls)^28^.

Case control data from the UK, the Netherlands, Uganda, Malawi and South Africa came from separate studies. The iGBS3 study which was embedded within the GBS3 study contributed the UK data. Cases and controls from the Netherlands were selected from the NoGBS study. The data from Uganda came from the ProGreSs birth cohort. We included published case-control data from South Africa^28^.

Across all studies, cases were defined as infants who developed iGBS disease during the first 90 days of life. Controls were infants who did not develop iGBS disease in the first 90 days of life, and were born to a woman who was colonised with GBS during pregnancy. Controls were serotype matched to cases in all countries, and additionally matched by gestational age at birth in South Africa^28^.

### Sample size

We calculated that a sample size of 150 cases of serotype III disease and three controls will be sufficient to estimate an 80% risk reduction (95% CI 68-87). The sample size for this study was based on the need for at least a 3:1 ratio of controls per case. Thus, ∼150 GBS cases of serotype III GBS will require matching with 450 controls (women colonised with serotype III) i.e. 450/0.328 ∼ 1370 women colonised with serotype III, which implies 1370/0.288 = 4800 women to be swabbed to ensure adequate women colonised with serotype III in the control group.

### Sample collection

#### Controls

Rectovaginal swabs were collected from pregnant women either at delivery or at 35-37 weeks gestation to detect maternal GBS colonisation. If GBS was detected, then informed consent was obtained to enrol the infant into the study at antenatal care visits or at delivery. In the Netherlands, rectovaginal swabs were collected from pregnant women after enrolment to the No GBS study. Cord blood samples were collected.

#### Cases

Cases were enrolled either at birth (prospectively) or at the time of disease (retrospectively). In South Africa both prospective and retrospective cases were identified through surveillance in national laboratories, with retrospective cases enrolled within 72 hours of laboratory confirmation of GBS disease. Whereas, in the UK, the Netherlands, Italy and Uganda, prospective cases were identified through GBS surveillance studies and retrospective cases identified through surveillance in national microbiology laboratories. When a GBS case was identified in a national laboratory the study team were notified and sought parental consent to enrol the infant in the study (unless already enrolled in the prospective cohort) and collect a serum sample (acute blood sample) from the infant and brief clinical and demographic details. With the exception of South Africa, all samples were shipped to the central laboratory at City St George’s, University of London for testing. Samples in South Africa were stored and analysed at the University of the Witswatersrand Vaccines and Infectious Disease Analytic Unit (Wits-VIDA)^28^. Details of the six studies are summarised in Table S1.

### Laboratory methods

GBS isolates were analysed using whole genome sequencing (WGS) to determine the carriage or disease serotype.

The standardised 6-plex anti-CPS IgG dLIA (GASTON assay) is a multiplex immunoassay based on Luminex MagPlex xMAP technology that allows for the measurement of GBS anti-CPS IgG (serotypes Ia, Ib, II, III, IV, and V) using six unique and spectrally distinct fluorescently dyed Luminex microsphere regions (one for each CPS; Pfizer Inc.) coupled to GBS CPS-PLL conjugates, as previously described^37^. Briefly, a 6-plex pool of microspheres was prepared using serotypes Ia, Ib, II, III, IV, and V conjugated to Poly-L-Lysine (PLL) (Pfizer Inc., New York, NY, USA) and coupled to Magplex^TM^ microspheres (Luminex, Austin, TX, USA) following the protocol described by Luminex Corporation^38^. CPS-PLL couplings were prepared at a working concentration of 10µg/mL.

Samples were diluted to 1/500, 1/5000, and 1/50,000 and incubated overnight with the prepared 6-plex pool of microspheres Each plate included an 11-point standard curve of multivalent vaccinee reference serum (Pfizer Inc, New York, NY, USA) diluted to 1/50 and serially diluted 2.5-fold, and two wells containing assay buffer acted as blank controls. The next day, the plates were washed with 100 µL/well of PBS/Tween (1xPBS/0.05%Tween/0.02%/Sodium Azide, pH 7.2) using a plate washer (Tecan Hydrospeed, Tecan, Reading, UK) with a magnetic base to retain the microspheres. Secondary antibody was added (R-Phycoerythrin Goat Anti-Human IgG Fcy specific, Jackson Laboratories 109-115-098, Jackson ImmunoResearch Ely, UK), at 1/500 (50 µL) to each well, and the plates were incubated for 90 (±15) minutes at room temperature under constant shaking (300 rpm).). Following the incubation, the plates were washed again, and 100 µL of wash buffer added to each well. The plates were read on a Bio-plex 200 (Bio-Rad Laboratories, Hercules, CA, USA) at high RP1 (high photomultiplier tube voltage). Median Fluorescent Intensity (MFI) values, generated from a minimum of 50 microspheres per region, were interpolated from the 11-point standard curve using log–log linear regression to derive IgG concentrations (µg/mL) for each sample.

#### Ethical approvals

Each of the six studies obtained their own ethical approvals as follows:

#### Netherlands

- Medisch Ethische Toetsingscommissie, NL63123.018.17

#### Italy

- Comitato Etico dell’Area Vasta Emilia Nord, 1079/2019/SPER/AOUMO – PREPARE

#### Malawi

- BAMBI: College of Medicine Research and Ethics Committee (COMREC), P.11/18/2536
- BAMBI II: College of Medicine Research and Ethics Committee (COMREC), P.01/19/2579

#### UK

- WP3: St George’s Research Ethics Committee, 2020.0221
- iGBS3: East Midlands - Derby Research Ethics Committee, REC red 20/EM/0260, IRAS ID 286733

#### Uganda

- Makerere University College of Health Sciences School of Medicine Research Ethics Committee (SOMREC), REC REF 2018-130
- Uganda National Council for Science and Technology (UNCST), HS 2496
- St George’s Research Ethics Committee, 2020.0024

### Statistical analysis

Demographics in cases and controls were summarised by country and overall. The number and percentage were presented for categorical data. Continuous variables were presented using the mean and standard deviation or median and range where appropriate.

Serotype-specific anti-CPS IgG concentrations below the lower limit of quantification (LLOQ) were imputed with half of the LLOQ. Anti-CPS IgG concentrations were log_10_ transformed for analysis. Geometric mean concentrations (GMC) were calculated as the anti-logarithm of the mean of the log-transformed values. Data were summarised by case-control status and by timing of disease onset (EOD or LOD).

The relative risk of disease was defined as the risk of disease relative to the predicted risk associated with the lowest antibody concentration in the data. In most cases this corresponds to half of the LLOQ. Relative risk curves were calculated using covariate adjust logit models (CALMs) ^32^ and the anti-CPS IgG levels associated with various levels of risk reduction were estimated from the relative risk curves.

The CALM method has been described in full elsewhere^32^. Briefly, the CALM is an extension of the scaled logit model^37^ and enables the inclusion of covariates in the analysis. The model estimates the probability of disease at specific antibody concentrations, i.e. P(iGBS | Antibody = a), and calculates relative risk to be the risk at a specific antibody concentration relative to the risk at the lowest antibody concentration as shown in Equation 1.

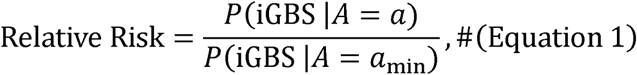

where *A* is the random antibody variable, *a* is the specific antibody concentration at which the relative risk is calculated, and *a*_min_ is the minimum antibody concentration for the assay. Assay LLOQs varied by serotype. In this analysis, we used the parametric implementation under the all-or-nothing assumption and models were adjusted for country where possible. In order to fit the model, at least 5 cases with antibody concentrations above the LLOQ were required and more than one case with antibody at or below the LLOQ.

The primary analysis included all infants with antibody data available from a cord blood sample or an acute blood sample collected within 7 days of disease onset. For analysis of EOD risk controls were serotype matched, such that all of the controls included in the analysis were born to women who were colonised with the same serotype as the cases. For the analysis of LOD risk, a mixture of homo- and heterotypic controls were used as in general only approximately 50% of LOD cases are born to women colonised with the same serotype^33^. Therefore, for LOD 50% of controls were randomly selected from those exposed to the serotype of interest and the other 50% were randomly selected from those exposed to a different serotype.

### Sensitivity analyses

A number of sensitivity analyses were conducted to assess the impact of different analysis decisions on the outcome and conclusions.

#### Analysis method

To allow for comparisons to be made between this study and published studies^28,31^, a sensitivity analysis was conducted using the Bayesian Weibull model, which has been described previously^39^. The Bayesian Weibull model predicts the absolute risk of disease at *or above* a specific antibody level, i.e. P(iGBS | Antibody ≥ a). From the absolute risk, the relative risk curves were generated in a similar way as in Equation 1. The key difference is that the cumulative probability is used to give the relative risk at *or <u>above</u>*a particular antibody level rather than the probability at a specific antibody as in the calculation using CALM. By generating relative risk curves using both the CALM and Bayesian Weibull model, we were able to compare the relative risk curves and SToRRs from both methods.

#### LOD controls

To assess the impact of the choice of controls for LOD, sensitivity analyses were conducted using 100% serotype matched controls instead of the mixture of controls used in the primary analysis. Relative risk curves and SToRRs were compared to the results from the primary analysis.

#### Antibody waning between birth and acute sample

All control infants had an antibody concentration measured from a cord blood sample. However, the majority of cases had an acute blood sample taken at the time of disease instead of at birth. Therefore, antibody concentrations for cases could be affected by antibody waning between birth and the time of disease. The half-life of naturally derived maternal anti-CPS IgG in infants has previously been estimated to be 27.4 (95% CI: 23.5, 32.9) days^40^. Given that LOD develops from one week to 3 months after birth, the antibody concentration in acute blood samples from LOD cases will have waned more than in samples from EOD cases.

We conducted a sensitivity analysis to investigate how potential antibody waning in acute samples can be accounted for in the analysis of LOD. Given that a large number of results were below the LLOQ, it was not possible to impute cord blood antibody concentrations from the individual antibody concentrations measured in acute samples. As an alternative, we used the following process to account for antibody waning in the analysis.

1. Use the distribution of ages at acute sample collection for LOD cases (Figure S1) to randomly assign an imputed “age” at acute sample to all controls and any cases with cord antibody data available.
2. Apply an imputed amount of antibody waning to the cord antibody concentrations using the random imputed “age” at sampling and an assumed antibody half-life of 27 days, as shown in Equation 2.

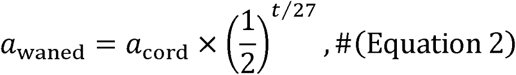

where a_waned_ is the imputed waned antibody, a_cord_ is the observed cord antibody, t is the imputed at acute sample, and 27 is the assumed antibody half-life.
3. Generate an adjusted relative risk curve using the imputed waned antibody concentrations for all cord blood samples, and the original antibody concentrations for acute blood samples.
4. Inflate the adjusted risk curve back to an assumed cord antibody level by translating the curve to the right using Equation 3 to increase all antibody values on the x-axis by the average amount of antibody decay associated with the mean age at acute blood sample collection.

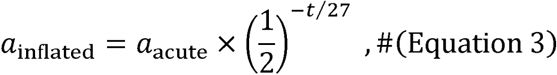

where *t* is the mean age at which acute blood samples were taken, and 27 is the assumed antibody half-life.

This process was repeated to generate 100 adjusted relative risk curves which were inflated back to an assumed cord antibody level. The median and 95% confidence intervals were estimated by the 2.5^th^, 50^th^ and 97.5^th^ percentiles.

#### Acute sample timing

For acute blood samples taken at the time of disease, there is the potential for antibody concentrations to change between disease onset and blood sample collection. This could be due to an early convalescent response to infection, causing an increase in antibody levels. Alternatively, antibody concentrations may decrease due to antibody waning or antibody consumption during infection. To assess the impact of the acute sampling window, three sensitivity analyses were conducted using samples collected within 3 days, 10 days, and all samples.

### Predicting vaccine efficacy

Predicted vaccine efficacy was computed based on the antibody risk curves. 1000 vaccine trials were simulated, with each trial consisting of 20,000 participants assigned 1:1 to receive a GBS vaccine or placebo. Serotype-specific cord blood antibody concentrations (*a_i,s_*) were simulated for infants, *i* = 1,2, …, 20,000 and serotypes *s* = Ia, III using antibody distributions derived from published GMCs and 95% confidence intervals (CI) for vaccine and placebo recipients from a were used to predict the risk of EOD (*p_i,s_*) and LOD (*q_i,s_*) for each infant using their individual phase 2 vaccine trial^28^. The serotype specific risk curves from the analysis of the PREPARE data antibody concentrations. Independent Bernoulli distributions were then used to assign the simulated disease status for each infant using their individual risk of disease. Predicted vaccine efficacy (pVE) was calculated for each simulated trial using Equation 4 and the median of all 1000 trials calculated, with the 95% confidence interval given by the 2.5^th^ and 97.5^th^ quantiles.

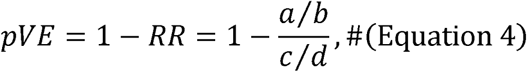

where *a* is the number of cases in the vaccine group, *b* is the total number in the vaccine group, *c* is the number of cases in the placebo group, and *d* is the total number in the placebo group.

## Data availability

Deidentified individual participant data and a data dictionary defining each field in the dataset will be made available for PREPARE WP3. The shared dataset will correspond to the data used for publication; raw data will not be provided. Additional related documents (including the study protocol, statistical analysis plan, and informed consent form) will not be publicly available but may be obtained from the corresponding author upon reasonable request. Data will be available from June 2026 via the City St George’s, University of London research data repository (https://sgul.figshare.com/). Data will be accessible for general research purposes without restriction.

Data from South Africa are available upon request, as per the data sharing statement for the Madhi 2023 publication (DOI: 10.1056/NEJMoa2116045).

## Acknowledgments

We thank the members of the NOGBS study groups (Supplement) and the technicians of Netherlands Reference Laboratory for Bacterial Meningitis for the collection of samples and characterisation of the group B streptococcal isolates, and the members of the GBS Italian Surveillance Network (Supplement) who actively collaborated in the collection of samples and clinical data.

## Author contributions

Conceptualisation: KLD, PTH, MWB, SAM

Data curation: KK, NT, MK

Formal analysis: LC

Funding acquisition: KLD, PTH, MWB, DvdB, MN, MV

Investigation: TH, EB, OD, CCK, SB

Project administration: MK, KK

Visualisation: LC

Writing – original draft: LC

Writing – review & editing: MV, KLD, AI, MWB, DvdB, KK, NA, SB, EB, AB, CCK, OD, NF, TH, AK, GK, MK, SAM, RM, FM, MN, NT, KFW, PTH

## Competing interests

PTH is a member of the UK Joint Committee on Vaccination and Immunisation (JCVI) and has received grant funding to his institution from the Bill & Melinda Gates Foundation, MinervaX, and Pfizer.

KLD and MV are members of the UK JCVI subcommittee on GBS vaccines.

MV received grant funding to her institution from the Gates Foundation

SAM institution received support for GBS sero-epidemiology studies from Gates Foundation, Pfizer, GSK and Minervax; and for GBS vaccine studies from Pfizer and PATH. The Immunisation and Vaccine Preventable Diseases Department at UKHSA has provided vaccine manufactures with post-marketing surveillance reports, which the Marketing Authorization Holders are required to submit to the UK Licensing authority in compliance with their Risk Management Strategy. A cost recovery charge is made for these reports.

## Supplementary Materials

Figs. S1 to S7 Tables S1 to S6

NOGBS Study Groups

GBS Italian Surveillance Network

## PREPARE WP3 Study Group

Phern Adams^16^, Adepeju Adedeji^17^, Hazel Alexander^18^, Sakina Ali^19^, Michelle Anderson^20^, Samya Armoush^21^, Naveen Athiraman^22^, Kavi Aucharaz^23^, Grace Audu^24^, Karen Austin-Smith^25^, Angela Ayuk^26^, Matthew Babirecki^27^, Kathryn Beardsall^28^, Samantha Beck^29^, "Sara Bennett^30^, Sumedha Bird^31^, Linda Bishop^32^, Isabel Bradley^33^, Nichola Brannen^34^, Helen Broomfield^35^, Matthijs C. Brouwer^14^, Anneka Burch^36^, Jane Cantliffe^37^, Lisa Capozzi^38^, Vineeth Cheruvalli^39^, Vivian Chu^20^, Paul Clarke^40^, Naomi Coleman^41^, Joanna Cook^36^, Chris Cooper^30^, Clinton Corin^42^, Roberta Creti^43^, Peter Cummins^44^, Laura Dean^45^, Jon Dorling^46^, Tomasz Dygas^47^, Sandra Dymond^48^, Cynthia Ebele^49^, Milli Edwards^41^, Louise Emmett^50^, Fabio Facchinetti^51^, Kirsty Farrington^52^, Shafqat Fatima^53^, Anam Fayadh^49^, Lynsey Felton^52^, Adam Finn^48^, Natasha Ford^20^, Lily French^55^, Clare Gallop^33^, Ramesh Ganapathy^50^, Emma Gardener^38^, Mandy Gill^56^, Nupur Goel^24^, Jenna Gould^29^, Sharon Gowans^52^, Laura Green^32^, Donna Griffiths^54^, Naomi Grimes^57^, Zoe Grindley^58^, Marleen A. Groenveld^14^, Julie Groombridge^22^, Suzanne Gunton^21^, Lara Robles Gutierrez^38^, Helen Harizaj^59^, Lauren Hatch^16^, Holly Hawkesford^31^, Victoria Hodgson^45^, Nicky Holland^60^, Stephanie Horridge^61^, Helen Hume^62^, Catriona Hussain^37^, Doris Iyamabo^19^, Lincy John^20^, Laura Johns^44^, Mark Johnson^46^, Stephen Jones^63^, Kelly Jukes^31^, Erum Khan^64^, Rizwan Khan^65^, Tahmina Khatun^66^, Amy Kitching^27^, Michelle Knight^50^, Athanasios (Thanos) Konstantinidis^67^, Rashmi Kuttysankaran^26^, Ivone Lancoma-Malcolm^66^, Katie Lang^16^, Alison Le Poidevin^68^, Sarah Lee^62^, Alex Li^33^, Emily Lifa^6^, Samantha Lissauer^6, 7, 12^, Titilope Majiyagbe^34^, Lumbani Makhaza^6^, Zoe Makin^30^, Khalid Mannan^65^, Sarah McCullough^69^, Rebekka Mccullough^69^, Joanna Mead^64^, Chiara Minotti^70, 71^, Hatem A. Mousa^53^, Baleke Munthali^6^, Richard Nicholl^49^, Jess Nutting^19^, Vita Nyasulu^6^, Sam Oddie^72^, Tanwa Ogbara^38^, Christina Oliver^63^, Chidera Nwokedi Onyeagor^38^, Nigel Osborne^44^, Prathiba Pai^36^, Leia Parry^57^, Santosh Pattnayak^59^, Stephane Paulus^73^, Daisy Pegler^30^, Jenny Pond^46^, Vennila Ponnusamy^60^, Catherine Postlethwaite^46^, Satodia Prakash^17^, Jennifer Pullen^63^, Eleanor Pyart^20^, Jyothi Rajeswary^56^, Shilpa Ratnaparkhi^34^, Natalie Reynolds^16^, Heidi Ribchester^32^, Chloe Rishton^69^, Joanna Robinson^35^, Grace Ryan^24^, Tabassum Safdar^58^, P Sashikumar^59^, Tim Scorer^68^, Chloe Scott^69^, Samantha Scott^40^, Jessica Sellick^63^, Fiona Shackley^21^, Julie Shaw^37^, Harriet Sheppard^34^, Ajay Sinha^66^, Ellen Smith^67^, Helen Smith^65^, Linde Snoek^74^, Nicola Squires^27^, Jasmine Stares^39^, Imogen Storey^41^, Louise Swaminathan^55^, Oluseun Tayo^54^, Simon Tazzyman^21^, Laura Thrasyvoulou^41^, Sangeeta Tiwary^26^, Chris Todd^67^, Lauren Trepte^24^, Serena Truocchio^75^, Lydia Ufton^55^, Merel N. van Kassel^76^, Ramamoorthy Varadan^39^, Stefania Vergnano^48^, Caterina Vocale^77^, Bryony Ward^73^, Tim Watts^35^, Sophie Webster^57^, Karen Wells^60^, Sonia White^25^, Eden Wildman^45^, Seren Wilson^31^, Vanessa Wong^28^, Julie Woollaston^52^, Priti Wuppalapati^78^, Cheryl Wyatt^61^, Edward Yates^42^, Tommaso Zini^79^

^16^Birmingham Women’s and Children’s NHS Foundation Trust. ^17^University Hospitals Coventry & Warwickshire NHS Trust. ^18^South Tees James Cook University Hospital. ^19^Luton & Dunstable University Hospital. ^20^Royal Free NHS Foundation Trust. ^21^Sheffield Children’s NHS Foundation Trust. ^22^Newcastle Upon Tyne. ^23^Barnsley Hospital NHS Foundation Trust. ^24^Chelsea and Westminster Hospital NHS Trust (West Middlesex University Hospital). ^25^Kettering General Hospital NHS Foundation Trust. ^26^Northumbria Healthcare Foundation Trust. ^27^Airedale NHS Foundation Trust. ^28^Cambridge University Hospitals NHS Foundation Trust /Addenbrookes. ^29^The Princess Alexandra Hospital NHS Trust. ^30^Stockport NHS Foundation Trust. ^31^South Warwickshire Foundation Trust. ^32^Southport and Ormskirk Hospital NHS Trust. ^33^Kingston Hospital. ^34^County Durham and Darlington NHS Foundation Trust. ^35^Guys and St Thomas NHS Foundation Trust. ^36^East Suffolk and North Essex NHS Foundation Trust. ^37^Nottingham University Hospitals. ^38^Lewisham and Greenwich NHS Trust. ^39^Northern Lincolnshire and Goole NHS Foundation Trust. ^40^Norfolk and Norwich University Hospitals NHS Foundation Trust. ^41^University Hospitals Birmingham. ^42^University Hospitals Sussex NHS Foundation Trust. ^43^Department of Infectious Diseases, Antibiotic Resistance and Special Pathogens Unit, Istituto Superiore di Sanità, Rome, Italy. ^44^Royal Devon and Exeter Foundation Trust Wonford Hospital. ^45^University Hospitals Plymouth NHS Trust. ^46^University Hospital Southampton NHS Trust. ^47^NHS Greater Glasgow and Clyde (Queen Elizabeth University Hospital). ^48^University Hospitals Bristol and Weston NHS Foundation Trust. ^49^London North West University Healthcare NHS Trust. ^50^Epsom and St Helier University Hospitals. ^51^University of Modena, Policlinico University Hospital, Modena, Italy. ^52^North Tees University Hospital. ^53^University Hospitals of Leicester. ^54^James Paget University Hospital. ^55^Maidstone and Tunbridge Wells NHS Trust. ^56^Sherwood Forest Hospitals NHS Foundation Trust. ^57^Kings College Hospital NHS Foundation Trust. ^58^Whiston (St Helens and Knowsley Teaching Hospitals NHS Trust). ^59^Medway NHS Foundation Trust. ^60^Ashford and St Peter’s. ^61^Lancashire Teaching Hospitals NHS Trust (LTHTR). ^62^Manchester University NHS Foundation Trust. ^63^Royal United Hospitals Bath. ^64^Milton Keynes Hospital NHS Trust. ^65^Barking, Havering And Redbridge University Hospitals NHS Trust. ^66^Barts Health NHS Trust. ^67^East Lancashire Teaching Hospitals NHS Trust. ^68^Portsmouth Hospital. ^69^Northern Care Alliance NHS Foundation Trust. ^70^Pediatric Research Center and Department of Neonatology. ^71^Department of Clinical Research, University Children’s Hospital Basel, Switzerland. ^72^Bradford Teaching Hospital. ^73^Oxford University Hospitals NHS Foundation Trust. ^74^Department of General Practice, Amsterdam UMC, University of Amsterdam, 1105 AZ, Amsterdam, the Netherlands. ^75^University of Modena, Italy. ^76^Department of Anaesthesiology, Amsterdam UMC, University of Amsterdam, 1105 AZ, Amsterdam, the Netherlands. ^77^Microbiology Unit, Ph.D., CRREM, IRCCS, S. Orsola University Hospital, Italy. ^78^Bolton NHS Foundation Trust. ^79^Pediatric Unit, Arcispedale Santa Maria Nuova, AUSL-IRCCS Reggio Emilia, Reggio Emilia, Italy.

