## Supplementary file for "Serological thresholds of risk reduction for infant group B streptococcus disease"

**The PDF file includes:**

Figs. S1 to S7

Tables S1 to S6

NOGBS Study Groups

GBS Italian Surveillance Network

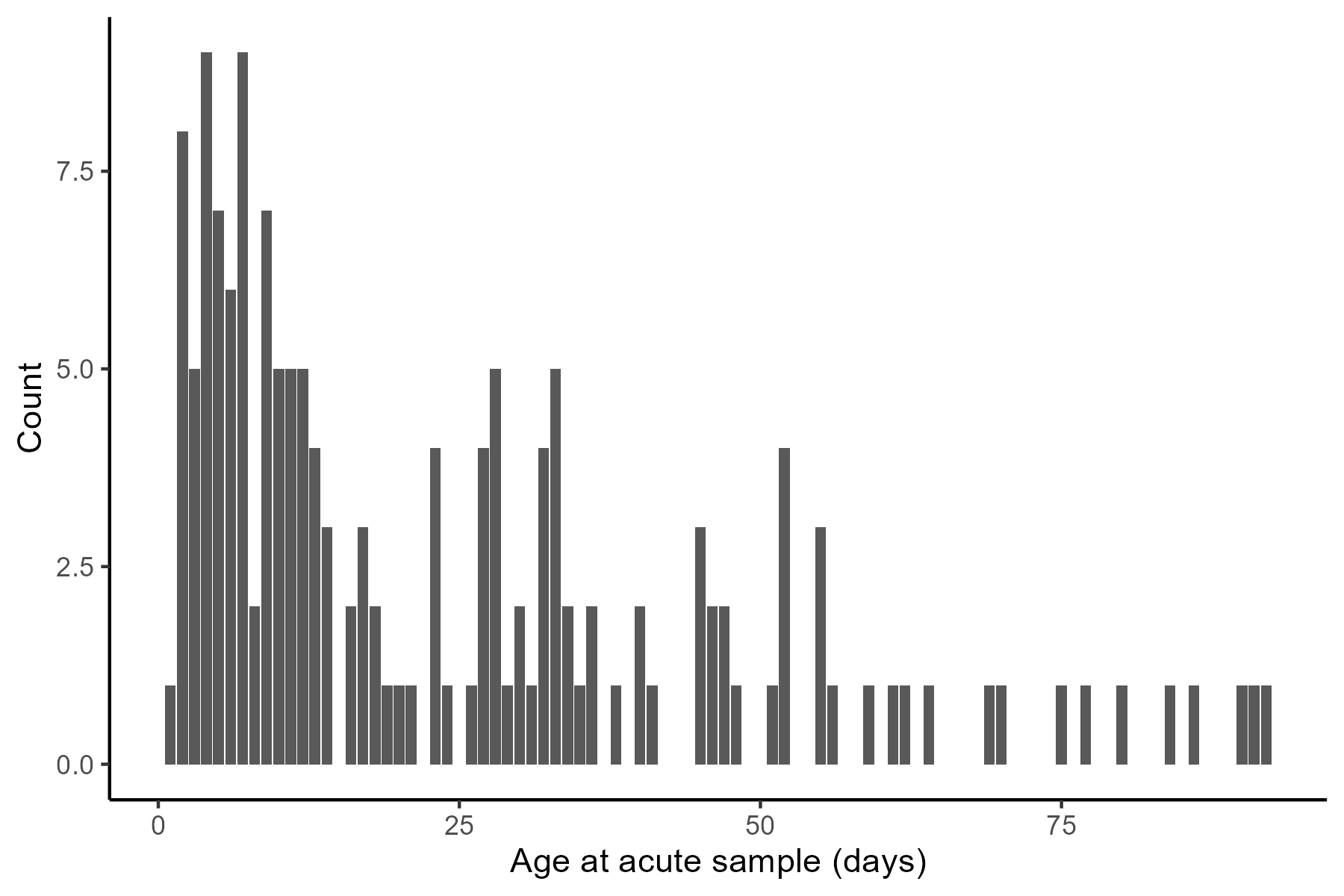

Fig. S1. Age at the time of acute blood sample collection (days) for serotype III cases. Bars show the number of acute blood samples collected at each age.

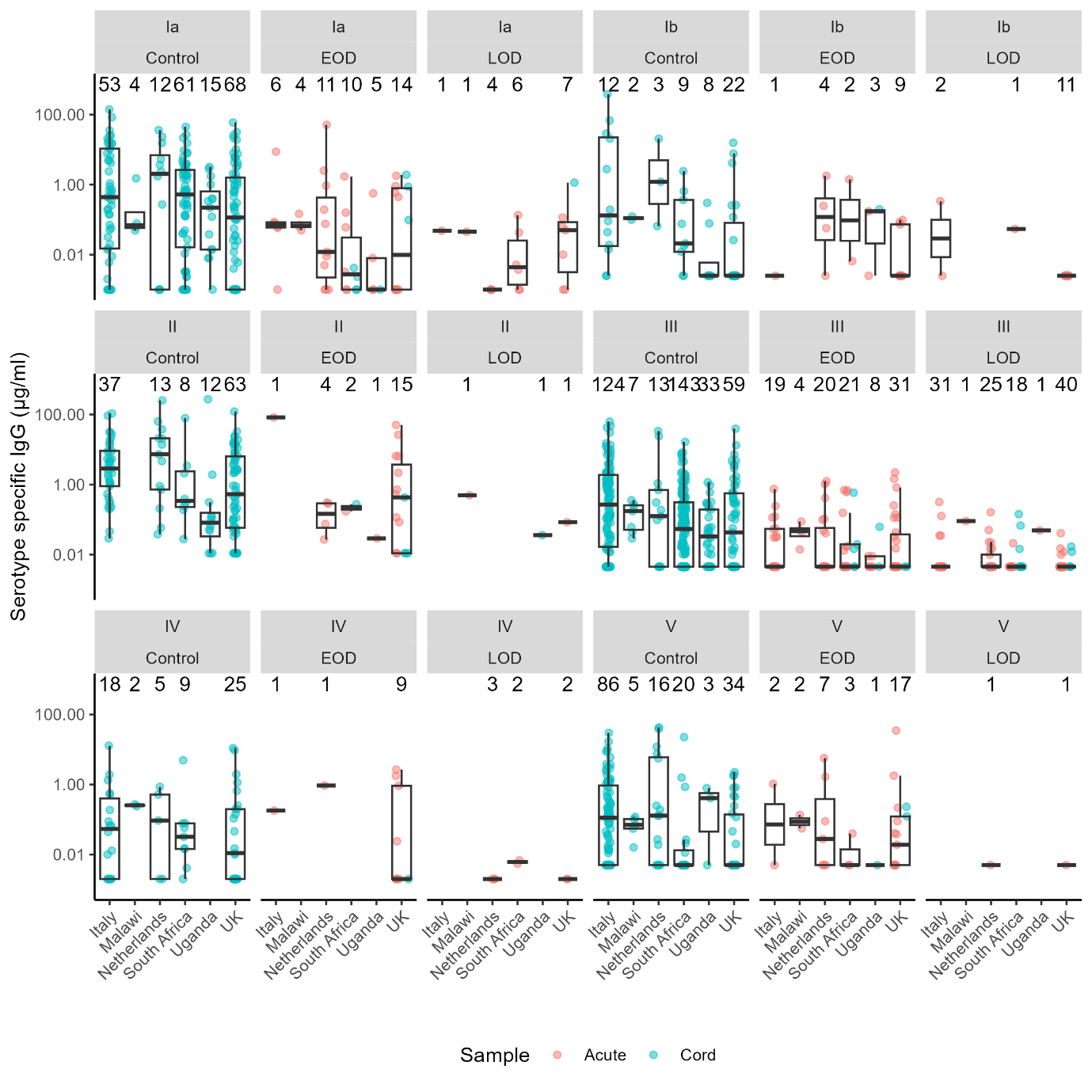
Fig. S2. Serotype-specific anti-capsular polysaccharide (anti-CPS) IgG concentrations (μg/ml) in controls, cases of early-onset disease (EOD; 0-6 days) and cases of late-onset disease (LOD; 7-89 days) broken down by country. Points show individual antibody concentrations in cord blood samples (blue) and acute blood samples (red), plotted on the log10 scale. Boxplots show the median and interquartile range in each country (x axis). The number of data points is annotated above each boxplot.

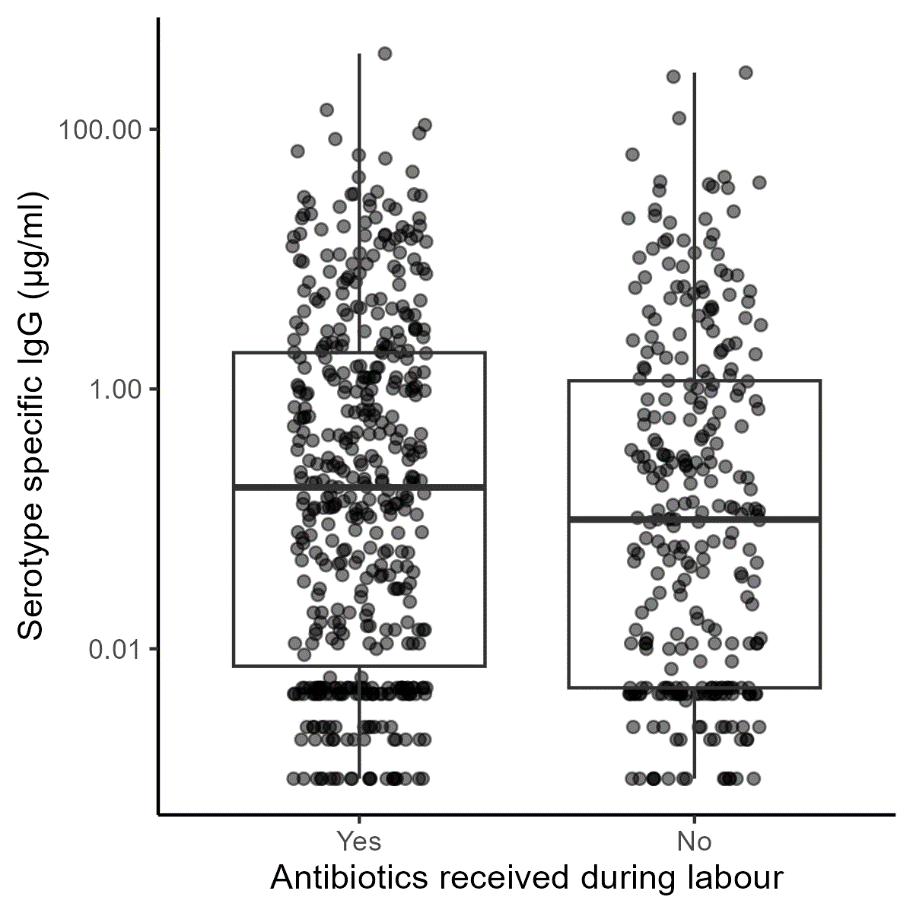

Fig. S3. Serotype specific anti-capsular polysaccharide IgG concentrations (μg/ml) in control infants, grouped by whether the infant’s mother received IAP during labour. Points show individual antibody concentrations plotted on the log10 scale. Boxplots show the median and interquartile range.

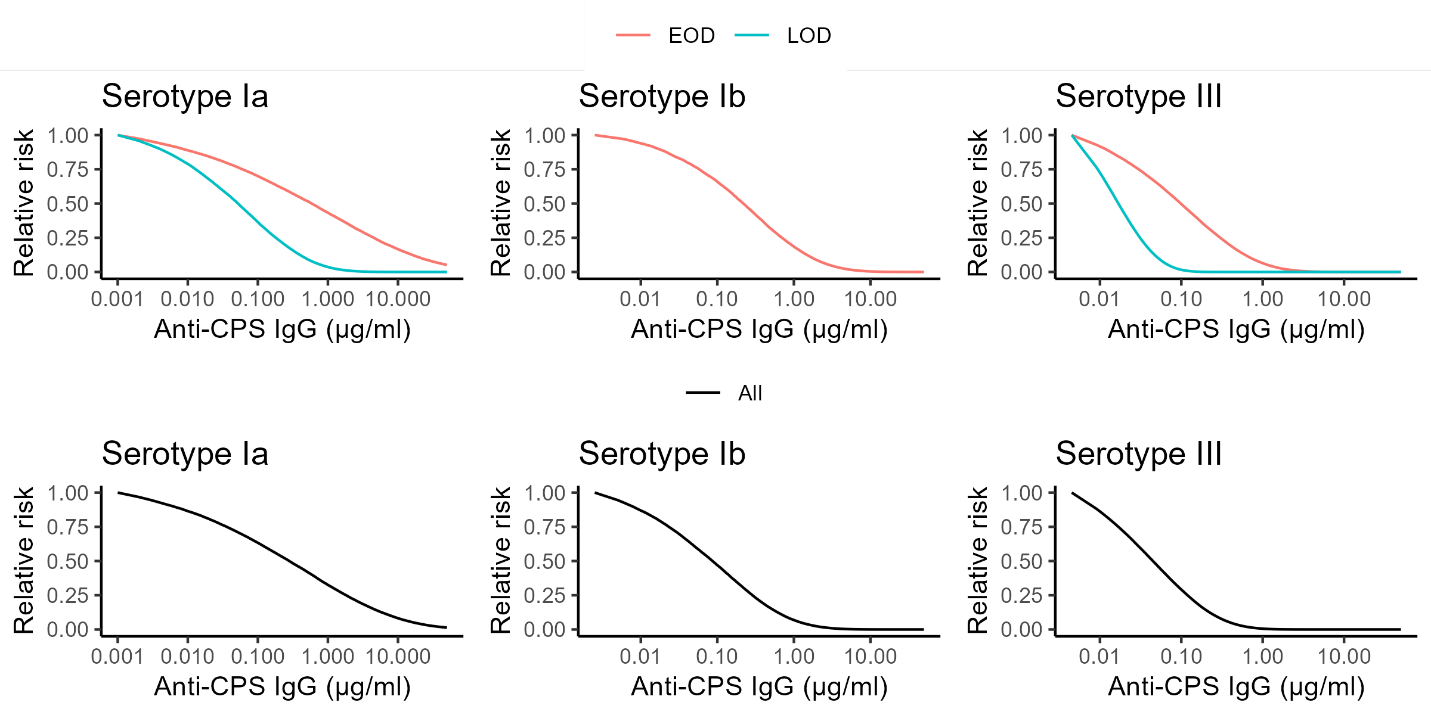

Fig. S4. Relative risk curves from the Weibull sensitivity analysis.

The relative risk curves for each serotype are shown in red for early-onset disease (EOD; 0-6 days), blue for late-onset disease (LOD; 7-89 days), and in black for EOD and LOD combined.

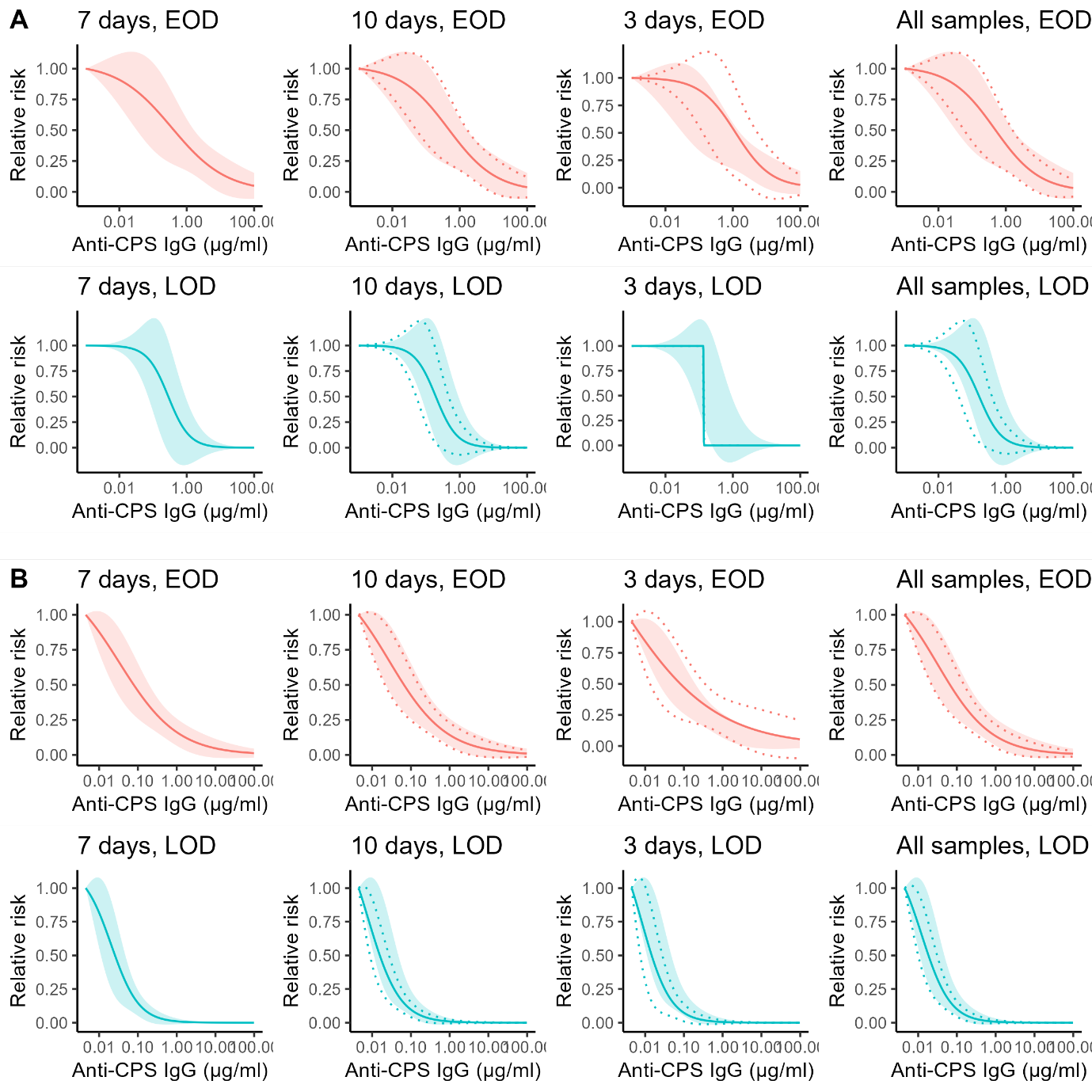

Fig. S5. Relative risk curves for the sample timing sensitivity analysis for A) serotype Ia and B) serotype III. Solid lines show the relative risk curves for the primary analysis (samples collected within 7 days of disease onset) and sensitivity analyses with various sampling windows (10 days, 3 days, all samples). The shaded region in all plots is the 95% confidence interval for the primary analysis (7 days). The 95% confidence intervals for the sensitivity analyses are represented by dotted lines. Red curves and confidence intervals show the analysis for early-onset disease (EOD; 0-6 days) and blue shows late-onset disease (LOD; 7-89 days)

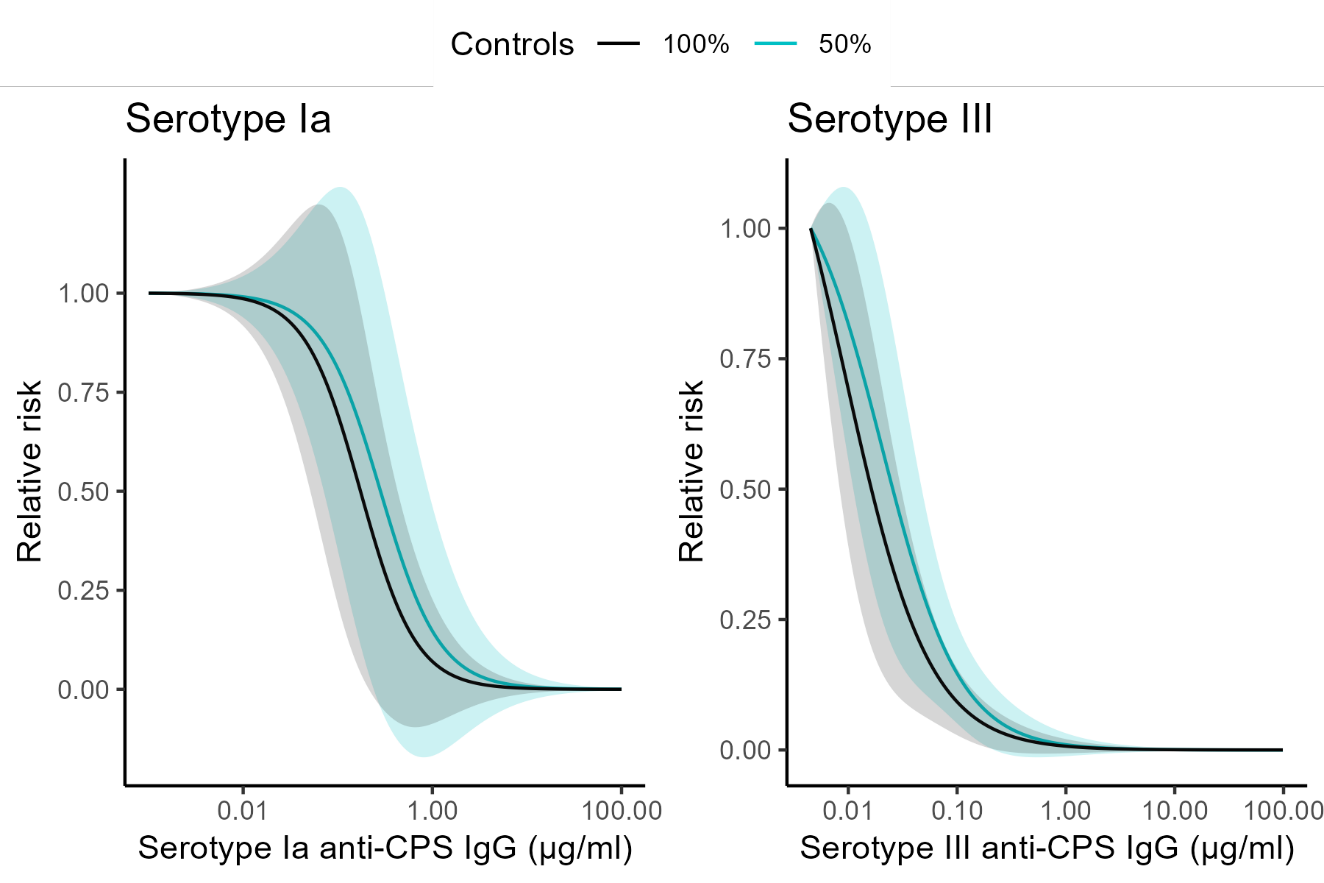

Fig. S6. Relative risk curves for the late-onset disease (LOD; 7-89 days) controls sensitivity analysis. Solid lines show the relative risk curves from the sensitivity analysis using 100% matched controls (black) and the primary analysis using 50% controls (blue). The 95% confidence intervals are shaded in blue for the primary analysis and grey for the sensitivity analysis.

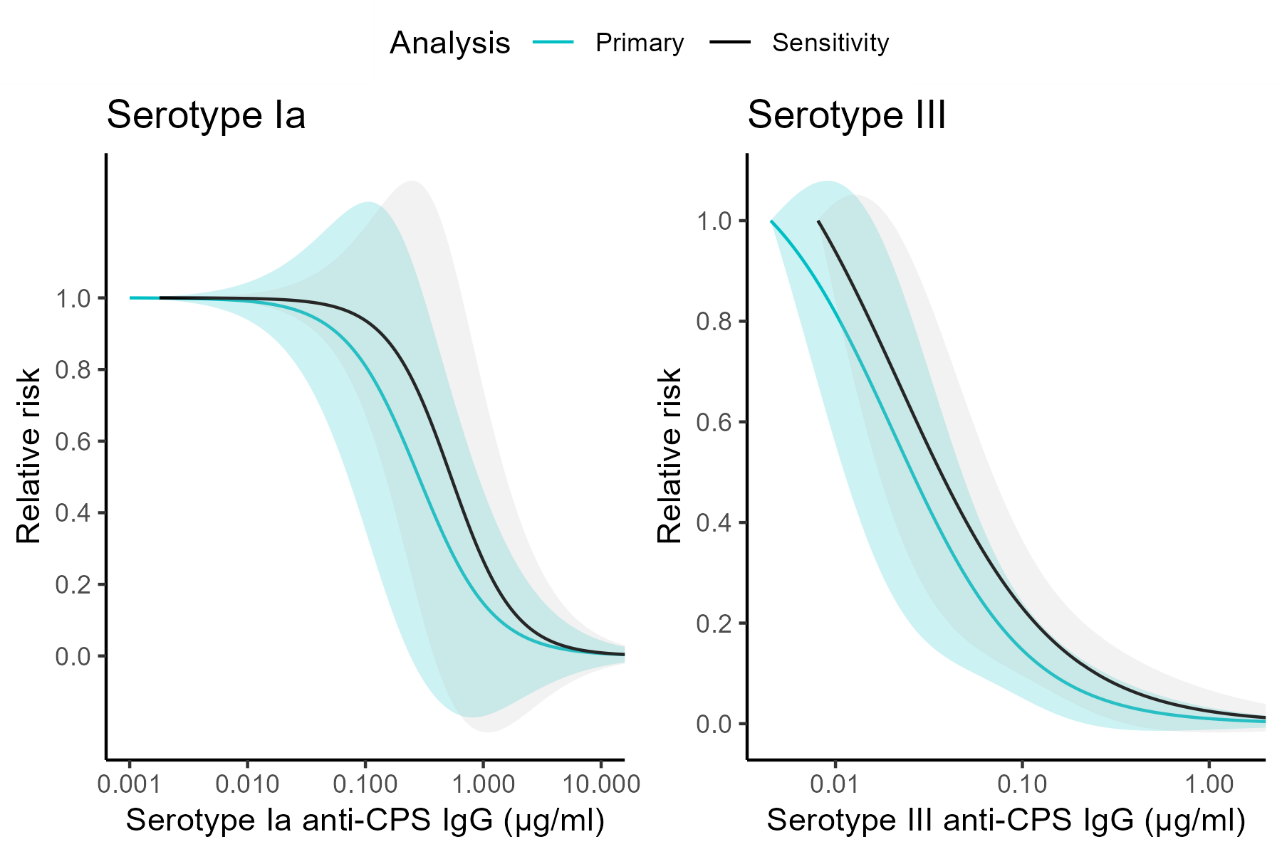

Fig. S7. Relative risk curves for the acute sample waning sensitivity analysis for serotypes Ia and III. The primary analysis curves and 95% confidence intervals are shown by the blue lines and shaded regions. The risk curves and 95% confidence intervals for the sensitivity analyses are shown by the black lines and grey shaded regions.

Table S1. Summary of study methods across sites

|  | **Italy** | **South Africa** | **Netherlands** | **Malawi** | **Uganda** | **UK** |
| --- | --- | --- | --- | --- | --- | --- |
| **Case definition** | 0-90 days, GBS positive from a usually sterile site. | Culture-confirmed GBS disease with identification of GBS from a normally sterile site | Culture-confirmed GBS disease with GBS identified from a usually sterile site (blood/CSF). | 0-90 days, GBS positive from a usually sterile site. | GBS Identified from a usually sterile site (blood/CSF) – Kawempe & Mulago Hospitals  0-90 days old. | 0-90 days of age, GBS identified from a normally sterile site. |
| **Case exclusions** | Unable to give written informed consent. | Refusal to consent to study participation  Receipt of any blood products in the past 4 weeks or anticipated during labour  Participant enrolled in any GBS vaccine trial | CSF drain in situ prior to development of meningitis | Mother ≤17 years of age  BW <1000g  GA < 28 weeks | Birth cohort: maternal age < 14 years  Active Surveillance: no written informed consent. | Birth cohort: <18 years of age, no written informed consent, not in study hospital.  Active Surveillance: no written informed consent. |
| **Case identification** | Active Surveillance | Prospective cases – identified through the cohort study  Retrospective cases – cases of GBS disease enrolled after GBS was identified | GBS identified by Netherlands Reference Laboratory or reported by treating physicians. | Active surveillance at QECH  BAMBI II study. | Active surveillance for GBS. Cases come from birth cohort enrolled at delivery (cord blood, maternal serum) and active surveillance (maternal and infant serum) | Active Surveillance – UK Health Security Agency, microbiology, paediatric & infection networks.  Other cases come from birth cohort enrolled at delivery. |
| **Control definition** |  | Infants born to mothers enrolled in the cohort study, who does not develop GBS disease in the first 90 days of life. The mother was colonised by a homologous serotype to a case to which the infant is matched | All pregnant women delivering in a participating hospital. For this manuscript only women colonized by GBS were selected |  | Cord blood from GBS swab positive controls that deliver term infants without GBS disease (serum from prospective cohort). | From GBS3 study |
| **Control exclusions** |  | Refusal to consent to study participation  Receipt of any blood products in the past 4 weeks or anticipated during labour  Participant enrolled in any GBS vaccine trial | Refusal to consent to study participation or refusal to share data with international partners | Mothers < 18 years of age  Newborn with severe life limiting congenital abnormality.  Normally resident outside Blantyre District. |  | See GBS3 study |
| **Control identification** | PREPARE WP3 Protocol | Cohort group (~20,000 mother-newborn dyads enrolled at or before delivery) | No GBS Part 2  Pregnant women expecting to deliver in a participating hospital | BAMBI I Study. | ProGreSs birth cohort | UK GBS3 birth cohort swab positive women at 35 weeks onwards. |
| **Matching** | Serotype | Serotype  Gestational age (34->37 weeks and ≥37 weeks) | Serotype | Serotype | Serotype | Serotype |
| **Study start and end date** | July 2020 – July 2022 | March 2019 – June 2020 | Jan 2018-current | March 2020-current | Feasibility study May 2019 – April 2020  Main Study May 2020-2022 | iGBS: 2019-2020  iGBS3: 2020-2022 |
| **Serology samples** | **Cases:**  Cord blood  Maternal serum  Infant serum | **Cases:**  Cord blood (cohort only)  Infant serum  Maternal blood (cohort only) | **Cases:**  Cord blood  Maternal serum  Infant serum  Infant DBS | **Cases:**  Cord blood  Maternal serum  Infant serum | **Cases:**  Cord blood (BC only)  Maternal serum  Infant serum | **Cases:**  Cord blood (BC only)  Maternal serum (AS only)  Infant serum (AS only)  Infant DBS |
|  | **Controls:**  Cord blood  Maternal serum | **Controls:**  Cord blood  Maternal blood | **Controls:**  Cord blood  Maternal serum | **Controls:**  Cord blood | **Controls:**  Cord blood  Maternal serum | **Controls:**  Cord blood |
| **Other samples** | **Cases:**  Maternal RVS | **Cases:**  Maternal RVS (cohort only) | **Cases:**  Maternal RVS | **Cases:**  Maternal RVS | **Cases:**  Maternal RVS | **Cases:**  Maternal RVS (BC only) |
|  | **Controls:**  Maternal RVS | **Controls:**  Maternal RVS | **Controls:**  Maternal RVS | **Controls:**  Maternal RVS | **Controls:**  Maternal RVS | **Controls:**  Maternal RVS |

AS: Active surveillance; BC: Birth cohort; BW: Birth weight; CSF: Cerebrospinal fluid; DBS: Dried blood spots; EOD: Early-onset disease (0-6 days); GA: Gestational age; LOD: Late-onset disease (7-89 days); RVS: Rectovaginal swab;

Table S2. Demographics

|  | **UK** | | **Netherlands** | | **Italy** | | **Uganda** | | **Malawi** | | **South Africa** | | **Overall** | |
| --- | --- | --- | --- | --- | --- | --- | --- | --- | --- | --- | --- | --- | --- | --- |
|  | **Case**  N = 192 | **Control**  N = 271 | **Case**  N = 115 | **Control**  N = 62 | **Case**  N = 67 | **Control**  N = 330 | **Case**  N = 24 | **Control**  N = 71 | **Case**  N = 13 | **Control**  N = 20 | **Case**  N = 77 | **Control**  N = 250 | **Case**  N = 488 | **Control**  N = 1,004 |
| Sample with antibody data available | | | | | | | | | | | | | | |
| Acute only | 173 (90%) | 0 (0%) | 115 (100%) | 0 (0%) | 67 (100%) | 0 (0%) | 17 (71%) | 0 (0%) | 13 (100%) | 0 (0%) | 57 (74%) | 0 (0%) | 442 (91%) | 0 (0%) |
| Cord and acute | 16 (8.3%) | 0 (0%) |  |  |  |  | 2 (8.3%) | 0 (0%) |  |  |  |  | 18 (3.7%) | 0 (0%) |
| Cord only | 3 (1.6%) | 271 (100%) | 0 (0%) | 62 (100%) | 0 (0%) | 330 (100%) | 5 (21%) | 71 (100%) | 0 (0%) | 20 (100%) | 20 (26%) | 250 (100%) | 28 (5.7%) | 1,004 (100%) |
| Serotype | | | | | | | | | | | | | | |
| Ia | 27 (14%) | 68 (25%) | 22 (19%) | 12 (19%) | 7 (10%) | 53 (16%) | 5 (21%) | 15 (21%) | 5 (38%) | 4 (20%) | 18 (23%) | 61 (24%) | 84 (17%) | 213 (21%) |
| Ib | 22 (11%) | 22 (8.1%) | 6 (5.2%) | 3 (4.8%) | 3 (4.5%) | 12 (3.6%) | 3 (13%) | 8 (11%) | 0 (0%) | 2 (10%) | 4 (5.2%) | 9 (3.6%) | 38 (7.8%) | 56 (5.6%) |
| II | 19 (9.9%) | 63 (23%) | 4 (3.5%) | 13 (21%) | 1 (1.5%) | 37 (11%) | 4 (17%) | 12 (17%) | 1 (7.7%) | 0 (0%) | 2 (2.6%) | 8 (3.2%) | 31 (6.4%) | 133 (13%) |
| III | 89 (46%) | 59 (22%) | 68 (59%) | 13 (21%) | 53 (79%) | 124 (38%) | 11 (46%) | 33 (46%) | 5 (38%) | 7 (35%) | 45 (58%) | 143 (57%) | 271 (56%) | 379 (38%) |
| IV | 14 (7.3%) | 25 (9.2%) | 5 (4.3%) | 5 (8.1%) | 1 (1.5%) | 18 (5.5%) | 0 (0%) | 0 (0%) | 0 (0%) | 2 (10%) | 3 (3.9%) | 9 (3.6%) | 23 (4.7%) | 59 (5.9%) |
| V | 21 (11%) | 34 (13%) | 10 (8.7%) | 16 (26%) | 2 (3.0%) | 86 (26%) | 1 (4.2%) | 3 (4.2%) | 2 (15%) | 5 (25%) | 5 (6.5%) | 20 (8.0%) | 41 (8.4%) | 164 (16%) |
| Infant sex | | | | | | | | | | | | | | |
| Female | 85 (45%) | 131 (48%) | 57 (50%) | 34 (55%) | 31 (46%) | 155 (47%) | 12 (50%) | 32 (45%) | 2 (15%) | 12 (63%) |  |  | 187 (46%) | 364 (48%) |
| Male | 106 (55%) | 140 (52%) | 58 (50%) | 28 (45%) | 36 (54%) | 175 (53%) | 12 (50%) | 39 (55%) | 11 (85%) | 7 (37%) |  |  | 223 (54%) | 389 (52%) |
| Unknown | 1 | 0 |  |  |  |  |  |  | 0 | 1 | 77 | 250 | 78 | 251 |
| Gestational age at birth (weeks) | | | | | | | | | | | | | | |
| <34 | 44 (23%) | 0 (0%) | 20 (17%) | 0 (0%) | 18 (27%) | 0 (0%) | 4 (29%) | 1 (1.4%) | 2 (15%) | 3 (15%) | 11 (14%) | 19 (7.6%) | 99 (21%) | 23 (2.3%) |
| 34 - <37 | 19 (9.9%) | 8 (3.0%) | 11 (9.6%) | 6 (9.7%) | 10 (15%) | 5 (1.5%) | 1 (7.1%) | 2 (2.8%) | 0 (0%) | 3 (15%) | 18 (24%) | 34 (14%) | 59 (12%) | 58 (5.8%) |
| ≥37 | 128 (67%) | 263 (97%) | 84 (73%) | 56 (90%) | 39 (58%) | 325 (98%) | 9 (64%) | 68 (96%) | 11 (85%) | 14 (70%) | 47 (62%) | 196 (79%) | 318 (67%) | 922 (92%) |
| Unknown | 1 | 0 |  |  |  |  | 10 | 0 |  |  | 1 | 1 | 12 | 1 |
| IAP received during labour | | | | | | | | | | | | | | |
| Yes | 31 (16%) | 157 (58%) | 13 (12%) | 10 (16%) | 27 (40%) | 285 (86%) | 0 (0%) | 0 (0%) | 4 (31%) | 11 (58%) | 0 (NA%) | 0 (NA%) | 75 (19%) | 463 (62%) |
| No | 160 (84%) | 114 (42%) | 98 (88%) | 52 (84%) | 40 (60%) | 45 (14%) | 21 (100%) | 70 (100%) | 9 (69%) | 8 (42%) | 0 (NA%) | 0 (NA%) | 328 (81%) | 289 (38%) |
| Unknown | 1 | 0 | 4 | 0 |  |  | 3 | 1 | 0 | 1 | 77 | 250 | 85 | 252 |
| Days between disease onset and acute sample collection | | | | | | | | | | | | | | |
| ≤3 days | 61 (33%) | 271 (100%) | 35 (30%) | 62 (100%) | 21 (49%) | 330 (100%) | 4 (17%) | 71 (100%) | 0 (NA%) | 20 (100%) | 47 (82%) | 250 (100%) | 168 (40%) | 1,004 (100%) |
| 4-7 days | 90 (49%) | 0 (0%) | 45 (39%) | 0 (0%) | 19 (44%) | 0 (0%) | 16 (67%) | 0 (0%) | 0 (NA%) | 0 (0%) | 7 (12%) | 0 (0%) | 177 (42%) | 0 (0%) |
| >7 days | 34 (18%) | 0 (0%) | 35 (30%) | 0 (0%) | 3 (7.0%) | 0 (0%) | 4 (17%) | 0 (0%) | 0 (NA%) | 0 (0%) | 3 (5.3%) | 0 (0%) | 79 (19%) | 0 (0%) |
| Unknown | 7 | 0 |  |  | 24 | 0 |  |  | 13 | 0 | 20 | 0 | 64 | 0 |
| Median (range) | 5 (0, 33) | 0 (0, 0) | 5 (0, 37) | 0 (0, 0) | 4 (0, 24) | 0 (0, 0) | 6 (0, 10) | 0 (0, 0) | NA (Inf, -Inf) | 0 (0, 0) | 1 (0, 9) | 0 (0, 0) | 4 (0, 37) | 0 (0, 0) |
| Median age at disease onset (range) | 1 (0, 87) | - | 1 (0, 86) | - | 12 (0, 85) | - | 1 (0, 23) | - | 1 (0, 60) | - | NA | - | 1 (0, 87) | - |
| Unknown | 1 | - | 0 | - | 0 | - | 0 | - | 0 | - | 77 | - | 78 | - |
| Disease onset | | | | | | | | | | | | | | |
| EOD | 111 (58%) | - | 66 (57%) | - | 31 (46%) | - | 21 (88%) | - | 10 (77%) | - | 38 (56%) | - | 277 (58%) | - |
| LOD | 80 (42%) | - | 49 (43%) | - | 36 (54%) | - | 3 (13%) | - | 3 (23%) | - | 30 (44%) | - | 201 (42%) | - |
| Unknown | 1 | - | 0 | - | 0 | - | 0 | - | 0 | - | 9 | - | 10 | - |

Unless specified otherwise, the table shows the number and proportion in each category.

* Infants were ineligible for inclusion in South Africa if their mother received IAP.

*EOD: Early-onset disease (0-6 days); IAP: Intrapartum antibiotics prophylaxis; LOD: Late-onset disease (7-89 days)*

Table S3. Serological thresholds of risk reduction (SToRR) from Bayesian Weibull analysis.

| **Serotype** | **Onset** | **75% SToRR** | **80% SToRR** | **90% SToRR** |
| --- | --- | --- | --- | --- |
| III | EOD | 0.301 | 0.388 | 0.726 |
|  | LOD | 0.032 | 0.036 | 0.053 |
|  | All | 0.12 | 0.15 | 0.262 |
| Ia | EOD | 4.513 | 7.062 | 21.845 |
|  | LOD | 0.181 | 0.242 | 0.484 |
|  | All | 1.824 | 2.777 | 7.933 |
| Ib | EOD | 0.701 | 0.919 | 1.787 |
|  | All | 0.288 | 0.378 | 0.744 |
| All | EOD | 3.51 | 5.246 | 13.834 |
|  | LOD | 0.04 | 0.049 | 0.082 |
|  | All | 0.758 | 1.091 | 2.659 |

EOD: Early-onset disease (0-6 days); LOD: Late-onset disease (7-89 days); SToRR: Serological threshold of risk reduction

Table S4. Serological thresholds of risk reduction (SToRR) from sensitivity analyses

| **Serotype** | **Onset** | **75% SToRR (95% CI)** | **80% SToRR (95% CI)** | **90% SToRR (95% CI)** |
| --- | --- | --- | --- | --- |
| **100% serotype matched controls for LOD** | | | | |
| Ia | LOD | 0.366 (0.115, 1.166) | 0.445 (0.133, 1.486) | 0.771 (0.178, 3.328) |
| III | LOD | 0.037 (0.018, 0.076) | 0.047 (0.024, 0.092) | 0.093 (0.049, 0.177) |
| **Sample timing – samples within 3 days of disease onset** | | | | |
| Ia | EOD | 4.608 (0.739, 28.726) | 6.53 (0.893, 47.738) | 17.452 (1.157, 263.25) |
|  | LOD | 0.137 (0.129, 0.146) | 0.137 (0.129, 0.146) | 0.138 (0.129, 0.146) |
| III | EOD | 0.876 (0.128, 6.006) | 1.795 (0.15, 21.461) | 15.431 (0.073, 3258.105) |
|  | LOD | 0.032 (0.015, 0.068) | 0.04 (0.02, 0.081) | 0.076 (0.038, 0.153) |
| **Sample timing – samples within 10 days of disease onset** | | | | |
| Ia | EOD | 2.893 (0.695, 12.036) | 4.618 (0.943, 22.603) | 17.327 (1.566, 191.762) |
|  | LOD | 0.429 (0.142, 1.302) | 0.528 (0.17, 1.639) | 0.949 (0.245, 3.673) |
| III | EOD | 0.332 (0.134, 0.82) | 0.522 (0.202, 1.35) | 1.938 (0.457, 8.215) |
|  | LOD | 0.037 (0.02, 0.067) | 0.047 (0.027, 0.083) | 0.097 (0.055, 0.172) |
| **Sample timing – no restriction on sample timing (all samples)** | | | | |
| Ia | EOD | 2.663 (0.719, 9.869) | 4.141 (0.983, 17.448) | 14.414 (1.683, 123.463) |
|  | LOD | 0.373 (0.126, 1.103) | 0.463 (0.154, 1.39) | 0.854 (0.228, 3.202) |
| III | EOD | 0.314 (0.133, 0.742) | 0.484 (0.197, 1.187) | 1.693 (0.45, 6.363) |
|  | LOD | 0.042 (0.024, 0.071) | 0.052 (0.031, 0.087) | 0.101 (0.06, 0.169) |
| **Acute sample waning** | | | | |
| Ia | LOD | 0.576 (0.129, 2.573) | 0.683 (0.145, 3.224) | 1.107 (0.18, 6.825) |
| III | LOD | 0.05 (0.026, 0.099) | 0.064 (0.034, 0.122) | 0.13 (0.061, 0.28) |

CI: Confidence interval; EOD: Early-onset disease (0-6 days); LOD: Late-onset disease (7-89 days); SToRR: Serological threshold of risk reduction

Table S5. The number of cases included in the analysis when the acute sampling window was varied.

| Serotype | Onset | 7 days (primary analysis) | 10 days | 3 days | All samples |
| --- | --- | --- | --- | --- | --- |
| Ia | EOD | 50 | 53 | 24 | 58 |
|  | LOD | 19 | 23 | 11 | 25 |
| III | EOD | 103 | 116 | 57 | 124 |
|  | LOD | 116 | 130 | 73 | 143 |

EOD: Early-onset disease (0-6 days); LOD: Late-onset disease (7-89 days)

Table S6. Predicted vaccine efficacy

| Serotype | Onset | Median number of placebo cases (range) | Median number of vaccine cases (range) | Median relative risk (95% CI) | Median vaccine efficacy (95% CI) |
| --- | --- | --- | --- | --- | --- |
| Ia | EOD | 56 (34, 79) | 9 (1, 22) | 0.17 (0.07, 0.31) | 83% (69, 93) |
|  | LOD | 11 (2, 22) | 0 (0, 3) | 0 (0, 0.14) | 100% (86, 100) |
| III | EOD | 72 (49, 99) | 18 (7, 36) | 0.25 (0.14, 0.41) | 75% (59, 86) |
|  | LOD | 40 (20, 61) | 4 (0, 12) | 0.09 (0.02, 0.22) | 91% (78, 98) |

CI: Confidence interval; EOD: Early-onset disease (0-6 days); LOD: Late-onset disease (7-89 days)

NOGBS Study Groups (Part I and II)

| **First name** | **Surname** | **Affiliation** |
| --- | --- | --- |
| Eveline | van Asbeck | Department of Obstetrics and Gynaecology, Tergooi Medical Centre, Hilversum, the Netherlands |
| Ron | van Beek | Department of Paediatrics, Amphia Hospital, Breda, the Netherlands |
| Vincent | Bekker | Department of Paediatrics, Leiden University Medical Centre, Leiden, the Netherlands |
| Maartje | van den Berg | Department of Paediatrics, Haaglanden Medical Centre, The Hague, the Netherlands |
| Geert Jan | Blok | Department of Neonatology, Northwest Clinics, Alkmaar, the Netherlands |
| Mijke | Breukels | Department of Paediatrics, Elkerliek Hospital, Helmond, the Netherlands |
| Alwin F.J. | Brouwer | Department of Paediatrics, Hospital of Nij Smellinghe, Drachten, the Netherlands |
| Renske | Cornelisse-van Vught | Department of Paediatrics, Canisius Wilhelmina Hospital, Nijmegen, the Netherlands |
| Luçan C. | Delemarre | Department of Paediatrics, Amstelland Hospital, Amstelveen, the Netherlands |
| Anouk | Dings | Department of Paediatrics, Gelre Hospitals, Apeldoorn, the Netherlands |
| Rienus A | Doedens | Department of Paediatrics, Martini Hospital, Groningen, the Netherlands |
| Stefan M. | van Dorth | Department of Paediatrics, Tjongerschans Hospital, Heerenveen, the Netherlands |
| Katja | de Graaff | Department of Obstetrics and Gynaecology, Reinier de Graaf Hospital, Delft, the Netherlands |
| Hester M. | Havers | Department of Paediatrics, Alrijne Hospital, Leiderdorp, the Netherlands |
| Jojanneke | Heidema | Department of Paediatrics, Antonius Hospital, Utrecht, the Netherlands |
| Marieke A.C. | Hemels | Department of Neonatology, Isala Clinics, Zwolle, the Netherlands |
| Maartje E.N. | van den Heuvel | Department of Paediatrics, OLVG, location West, Amsterdam, the Netherlands |
| Marion | van Hoorn | Department of Obstetrics and Gynaecology, Haga Hospital, The Hague, the Netherlands |
| Marlies A. | van Houten | Department of Infectious Diseases (LUCID), Leiden University Medical Centre, Leiden, the Netherlands |
| Monique A.M. | Jacobs | Department of Paediatrics, Slingeland Hospital, Doetinchem, the Netherlands |
| Arieke | Janse | Department of Paediatrics, Gelderse Vallei Hospital, Ede, the Netherlands |
| Miranda | de Jong | Department of Paediatrics, Albert Schweitzer Hospital, Dordrecht, the Netherlands |
| Anton H. | van Kaam | Department of Neonatology, Emma Children’s Hospital, Amsterdam UMC, Amsterdam, the Netherlands |
| Ageeth | Kaspers | Department of Paediatrics, Medisch Spectrum Twente, Twente, the Netherlands |
| Anne A.M.W. | van Kempen | Department of Paediatrics, OLVG, location East, Amsterdam, the Netherlands |
| Kristine | Klúčovská | Department of Paediatrics, Treant Hospital Group, Hoogeveen, the Netherlands |
| Karen | Korbeek | Department of Paediatrics, St. Jansdal Hospital, Harderwijk, the Netherlands |
| René F. | Kornelisse | Department of Paediatrics, Erasmus Medical Centre, Rotterdam, the Netherlands |
| Anke G. | Kuijpers | Department of Paediatrics, Bernhoven Hospital, Uden, the Netherlands |
| Taco W. | Kuijpers | Department of Peadiatrics, Emma Children’s Hospital, Amsterdam UMC, Amsterdam, the Netherlands |
| Elizabeth | van Leeuwen | Department of Obstetrics and Gynaecology, Amsterdam UMC, Amsterdam, the Netherlands |
| Jeannette | von Lindern | Department of Paediatrics, Groene Hart Hospital, Gouda, the Netherlands |
| Carmen | Lorente Flores | Department of Neonatology, Maxima Medical Centre, Veldhoven, the Netherlands |
| Wing Kit | Man | Department of Neurology, Amsterdam UMC, University of Amsterdam, the Netherlands |
| Karen | Van Mechelen | Department of Neonatology, Maastricht University Medical Centre, Maastricht, the Netherlands |
| Clemens B. | Meijssen | Department of Paediatrics, Meander Medical Centre, Amersfoort, the Netherlands |
| Jeroen | Noordzij | Department of Paediatrics, Reinier de Graaf Hospital, Delft, the Netherlands |
| Annemarie | Oudshoorn | Department of Paediatrics, Gelre Hospitals, Apeldoorn, the Netherlands |
| Frans B. | Plötz | Department of Paediatrics, Tergooi Medical Centre, Hilversum, the Netherlands |
| Marjolijn | Quaak | Department of Paediatrics, Dijklander Hospital, Hoorn, the Netherlands |
| Maaike | van Rossem | Department of Paediatrics, Rijnstate Hospital, Arnhem, the Netherlands |
| Maarten | Rijpert | Department of Paediatrics, Zaans Medical Centre, Zaandam, the Netherlands |
| Machteld A.G. | van Scherpenzeel-de Vries | Department of Paediatrics, Frisius Medical Centre, Leeuwarden, the Netherlands |
| Irene | Schiering | Department of Paediatrics, Spaarne Gasthuis, Haarlem, the Netherlands |
| George | Shabo | Department of Paediatrics, Hospital Group Twente, Twente, the Netherlands |
| Nina M. | van Sorge | Netherlands Reference Laboratory for Bacterial Meningitis, Amsterdam UMC Location AMC, Amsterdam, Netherlands |
| Jacqueline U.M. | Termote | Department of Neonatology, University Medical Centre Utrecht, Utrecht, the Netherlands |
| Gerdien A. | Tramper-Stranders | Department of Paediatrics, Franciscus Gasthuis, Rotterdam, the Netherlands |
| Mirjam | van Veen | Department of Paediatrics, Haga Hospital, The Hague, the Netherlands |
| Joost | van de Ven | Department of Obstetrics and Gynaecology, Elkerliek Hospital, Helmond, the Netherlands |
| Marlies | Vermaas | Department of Paediatrics, Admiraal de Ruyter Hospital, Goes, the Netherlands |
| Marjoke | Verweij | Department of Paediatrics, Viecuri Medical Centre, Venlo, the Netherlands |
| Douwe H. | Visser | Department of Neonatology, Emma Children’s Hospital, Amsterdam UMC, Amsterdam, the Netherlands |
| Karlijn C. | Vollebregt | Department of Obstetrics and Gynaecology, Spaarne Gasthuis, Haarlem, the Netherlands |
| Wouter J. | de Waal | Department of Paediatrics, Diakonesse Hospital, Utrecht, the Netherlands |
| Anne-Marie | van Wermeskerken | Department of Paediatrics, Flevohospital, Almere, the Netherlands |
| Janneke F. | Wilms | Department of Paediatrics, BovenIJ Hospital, Amsterdam, the Netherlands |
| Tom F.W | Wolfs | Department of Paediatrics, University Medical Centre Utrecht, Utrecht, the Netherlands |
| Maurice G.A.J. | Wouters | Department of Obstetrics and gynaecology, Amsterdam UM, Amsterdam, the Netherlands |
| Angela C.M. | van Zijl | Department of Neonatology, University Medical Centre Utrecht, Utrecht, the Netherlands |

GBS Italian Surveillance Network

| **Name** | **Surname** |  |  |
| --- | --- | --- | --- |
| Barbara | Perrone | Neonatal Intensive Care Unit | United Hospitals of Ancona, Italy |
| Giuseppe | Latorre | Neonatal Intensive Care Unit | Miulli Hospital, Acquaviva delle Fonti, Italy |
| Matilde | Ciccia | Neonatal Intensive Care Unit | Maggiore Hospital, Bologna, Italy |
| Sofia | Spinedi | Neonatal Intensive Care Unit | Ospedale Maggiore, Bologna, Italy |
| Morena | De Angelis | Neonatal Intensive Care Unit | Maggiore Hospital, Bologna, Italy |
| Simone | Ambretti | Microbiology Laboratory | IRCCS, S. Orsola Policlinic, Italy |
| Arianna | Dondi | Pediatric Emergency Unit | IRCCS, S. Orsola Policlinic, Italy |
| Marcello | Lanari | Pediatric Emergency Unit | IRCCS, S. Orsola Policlinic, Italy |
| Silvia | Fanaro | Neonatal Intensive Care Unit | Sant’Anna Hospital, Ferrara, Italy |
| Claudia | Venturelli | Microbiology Laboratory | University Hospital Policlinico, Modena, Italy |
| Filippo | Ferrari | Microbiology Laboratory | University Hospital Policlinico, Modena, Italy |
| Martina | Buttera | Neonatal Intensive Care Unit | University Hospital Policlinico, Modena, Italy |
| Valeria | Capone | Neonatal Intensive Care Unit | University Hospital Policlinico, Modena, Italy |
| Riccardo | Barberini | Neonatal Intensive Care Unit | University Hospital Policlinico, Modena, Italy |
| Mattia | Iaccheri | Obstetrics and Gynecology Clinic | University Hospital Policlinico, Modena, Italy |
| Sara | Ovani | Obstetrics and Gynecology Clinic | University Hospital Policlinico, Modena, Italy |
| Fabio | Facchinetti | Obstetrics and Gynecology Clinic | University Hospital Policlinico, Modena, Italy |
| Daniela | Menichini | Obstetrics and Gynecology Clinic | University Hospital Policlinico, Modena, Italia |
| Lucia | Fidanza | Clinical Analysis Laboratory | Baggiovara Hospital, Modena, Italy |
| Lucia | Marrozzini | Pediatric Unit | Ramazzini Hospital, Carpi, Italy |
| Tommaso | Zini | Pediatric Unit | Santa Maria Nuova Archhospital, AUSL-IRCCS Reggio Emilia, Italy |
| Lorenza | Baroni | Neonatal Intensive Care Unit | Santa Maria Nuova Archhospital, AUSL-IRCCS Reggio Emilia, Italy |
| Belinda | Benenati | Pediatric Unit | Guglielmo da Saliceto Hospital, Piacenza, Italy |
| Giacomo | Biasucci | 1. Pediatric and Neonatal Unit 2. Medical and Surgical Department | Guglielmo da Saliceto Hospital, Piacenza, Italy |
| Federico | Marchetti | Pediatric and Neonatal Unit | Santa Maria delle Croci Hospital, Ravenna, Italy |
| Giancarlo | Piccinini | Pediatric and Neonatal Unit | Santa Maria delle Croci Hospital, Ravenna, Italy |
| Lucia | Gambini | Neonatal Intensive Care Unit | University Hospital Policlinico, Parma, Italy |
| Carlo | Ferrari | Microbiology Laboratory | University Hospital Policlinico, Parma, Italy |
| Cristina | Tuoni | Neonatal Intensive Care Unit | University Hospital Company,Pisa, Italy |
| Marcello | Stella | Neonatal Intensive Care Unit | Bufalini Hospital, Cesena, Italy |
| Monica | Cricca | Microbiology Laboratory | Pieve Sestina Laboratory, Italy |
| Maria Chiara | China | Neonatal Intensive Care Unit | Infermi Hospital, Rimini, Italy |
| Irene | Papa | Neonatal Intensive Care Unit | Infermi Hospital, Rimini, Italy |
| Eleonora | Scapillati | Neonatal Intensive Care Unit | Fatebenefratelli Hospital, Roma, Italy |
| Cristina | Haas | Neonatal Intensive Care Unit | Fatebenefratelli Hospital, Roma, Italy |
| Cinzia | Auriti | Neonatal Intensive Care Unit | Bambin Gesù Pediatric Hospital, Roma, Italy |
| Jenny | Bua | Neonatal Intensive Care Unit | IRCCS Burlo Garofalo, Trieste, Italy |
| Nicola | Laforgia | Neonatal Intensive Care Unit | University Hospital Policlinico, Bari, Italy |
| Sabrina | Loprieno | Neonatal Intensive Care Unit | University Hospital Policlinico, Bari, Italy |
| Gianfranco | Maffei | Neonatal Intensive Care Unit | United Hospitals, Foggia, Italy |
| Angela | Lanzoni | Pediatric Unit | Santa Maria della Scaletta Hospital, Imola, Italy |
